# Phase I/II Trial of Brogidirsen: Dual-Targeting Antisense Oligonucleotides for Exon 44 Skipping in Duchenne Muscular Dystrophy

**DOI:** 10.1101/2024.08.28.24312624

**Authors:** Hirofumi Komaki, Eri Takeshita, Katsuhiko Kunitake, Takami Ishizuka, Yuko Shimizu-Motohashi, Akihiko Ishiyama, Masayuki Sasaki, Chihiro Yonee, Shinsuke Maruyama, Eisuke Hida, Yoshitsugu Aoki

## Abstract

Duchenne muscular dystrophy (DMD) is a severe muscle disorder caused by mutations in the DMD gene, resulting in dystrophin loss. Exon-skipping using antisense oligonucleotides (ASO) is a promising approach that partially restores dystrophin by correcting the frameshift during pre-mRNA splicing. However, a weakness of the current approach is that it is mutation-specific and has poor efficacy. To address these, we aim to develop brogidirsen, a new dual-targeting ASO that targets two sequences in exon 44 of the *DMD* using phosphorodiamidate morpholino oligomer. Here, we conducted an open-label, dose-escalation, Phase I/II trial to evaluate the safety, pharmacokinetics, and activity of brogidirsen, administered intravenously to six ambulant patients with DMD amenable to exon 44 skipping. The study consisted of a dose-escalation part to determine the optimal doses, followed by extended treatment with 40 mg/kg or 80 mg/kg weekly dose for 24 weeks. There were no serious adverse events related to brogidirsen. The results indicated a dose-dependent increase in dystrophin levels, reaching 10.27% and 15.79% of the normal level in the two cohorts. Motor functional tests suggested a trend toward maintaining or slightly improving motor function. There was a dose-dependent increase in Cmax and AUC0–t. High-throughput proteomic assays revealed that serum proteins such as PADI2, TTN, MYOM2, and MYLPF were observed to reduce, suggesting them as biomarkers for therapeutic effects. Notably, in vitro assays using urine-derived cells from patients with DMD support brogidirsen’s high efficacy in the first-in-human studies. These promising results warrant a subsequent multinational trial for DMD.

## Introduction

Duchenne muscular dystrophy (DMD) is a fatal inherited muscle disease caused by mutations in the DMD gene located on the short arm of the X chromosome (Xp21.2). DMD gene encodes dystrophin, a broad membrane integrator stabilising skeletal muscle myofibers’ plasma membrane.^1,2,3^ DMD almost exclusively affects males and has a global prevalence of approximately 19.8 in 100,000 live male births (i.e., approximately one in every 5,000 births).^4^ The initial symptoms, including frequent falls, are observed around 3 years of age.^5^ The associated muscle weakness progressively worsens over time. Typically, patients with DMD would be unable to walk and require a wheelchair by 8–14 years of age.^5^ However, no definitive treatments are currently for preventing muscle degeneration and necrosis.

Novel therapies for DMD are under investigation.^6,7^ Significant progress has been made in exon-skipping therapy, which shifts an out-of-frame deletion to an in-frame. This is accomplished using short, single-stranded antisense oligonucleotides (ASOs), approximately 20–30 bases in length, to sterically inhibit the complementary sequence of the target pre-mRN, thus regulating gene splicing.^8^ Conceptually, exon-skipping therapy for DMD is expected to improve muscle function by facilitating the expression of short but functional dystrophin proteins. Approximately 80% of patients with DMD, including those with deletions and duplication mutations in the DMD gene, can be treated with exon-skipping therapy.^9,10,11,12,13^ Preclinical studies in animal models of DMD have demonstrated restoring functional dystrophin and suppressing symptom progression with exon-skipping therapy.^14,15,16,17,18,19,20^ These promising findings have led to the initiation of clinical trials on exon-skipping treatment involving patients with DMD.

Approximately 60% of patients with DMD harbor a deletion in one or more exons; deletions cluster between exons 45 and 55.^21,22^ Therefore, ASO formulations targeting exons within this region were the first to be developed. Several ASOs targeting this region have been approved by the US Food and Drug Administration (FDA) to treat patients with DMD.^23^ These include the conditional approval of eteplirsen targeting exon 51,^24,25,26^ the approval of golodirsen targeting exon 53,^27,28,29^ and the approval of viltolarsen targeting exon 53 (NS-065/NCNP-01) in both the US and Japan.^30,31,32^ All ASOs currently approved for DMD treatment are phosphorodiamidate morpholino oligonucleotides (PMOs). They are water-soluble because of their morpholine ring structure. They bind strongly to target mRNA precursors, do not induce a Toll-like receptor-mediated immune response, have nuclease resistance *in vivo*, and preferentially reach regenerating myotubes.^33^

Despite promising findings for these ASOs, an unmet need remains for patients with deletions in other exons.^18,34,35^ We developed NS-089/NCNP-02, brogidirsen, as the first dual-targeting ASO_-_based exon 44-skipping drug.^36^ Brogidirsen is inherently safe and was developed using a novel high-activity sequence discovery method with a sequence-linked ASO that imparts a high level of exon-skipping activity.^36^ Patients amenable to exon 44 skipping harbour the following mutation patterns: deletions in DMD exons 14–43, 19–43, 30–43, 35–43, 36–43, 40–43, 42–43, 45, and 45–54.^9,34^ It is estimated that 6.2% of all patients with DMD will benefit from this skipping.^9^

This first-in-human phase I/II study evaluated the safety, efficacy, and pharmacokinetics of brogidirsen in six patients with DMD and a DMD mutation amenable to exon 44 skipping. Additionally, we evaluated, for the first time, the usefulness of MYOD1-transduced urine-derived cells (MYOD1-UDCs) in assessing the efficacy of brogidirsen in a DMD clinical trial.^37,38^

## RESULTS

### Patients

Between December 2, 2019, and October 19, 2021, six patients from NCNP Hospital in Japan aged 4–13 years were enrolled. All study visits were conducted at the NCNP, apart from Part 2 (except for motor function assessment) for a single patient at Kagoshima University Hospital (Kagoshima). One and five patients were DMD exon 45–54- and 45-deletion, respectively; all were ambulant, and all but one was being treated with corticosteroids (**Table 1**). All patients participated in Parts 1 and 2 of the trial. The cumulative dose of brogidirsen during treatment ranged from 50 to 560 mg in Part 1 and 19,560 to 62,520 mg in Part 2 (**Table 1**). Based on the data collected in Part 1, 40 mg/kg and 80 mg/kg weekly brogidirsen doses were used in Part 2 (Cohort 1 and 2, respectively), with three patients included in each cohort.

**Table 1:**
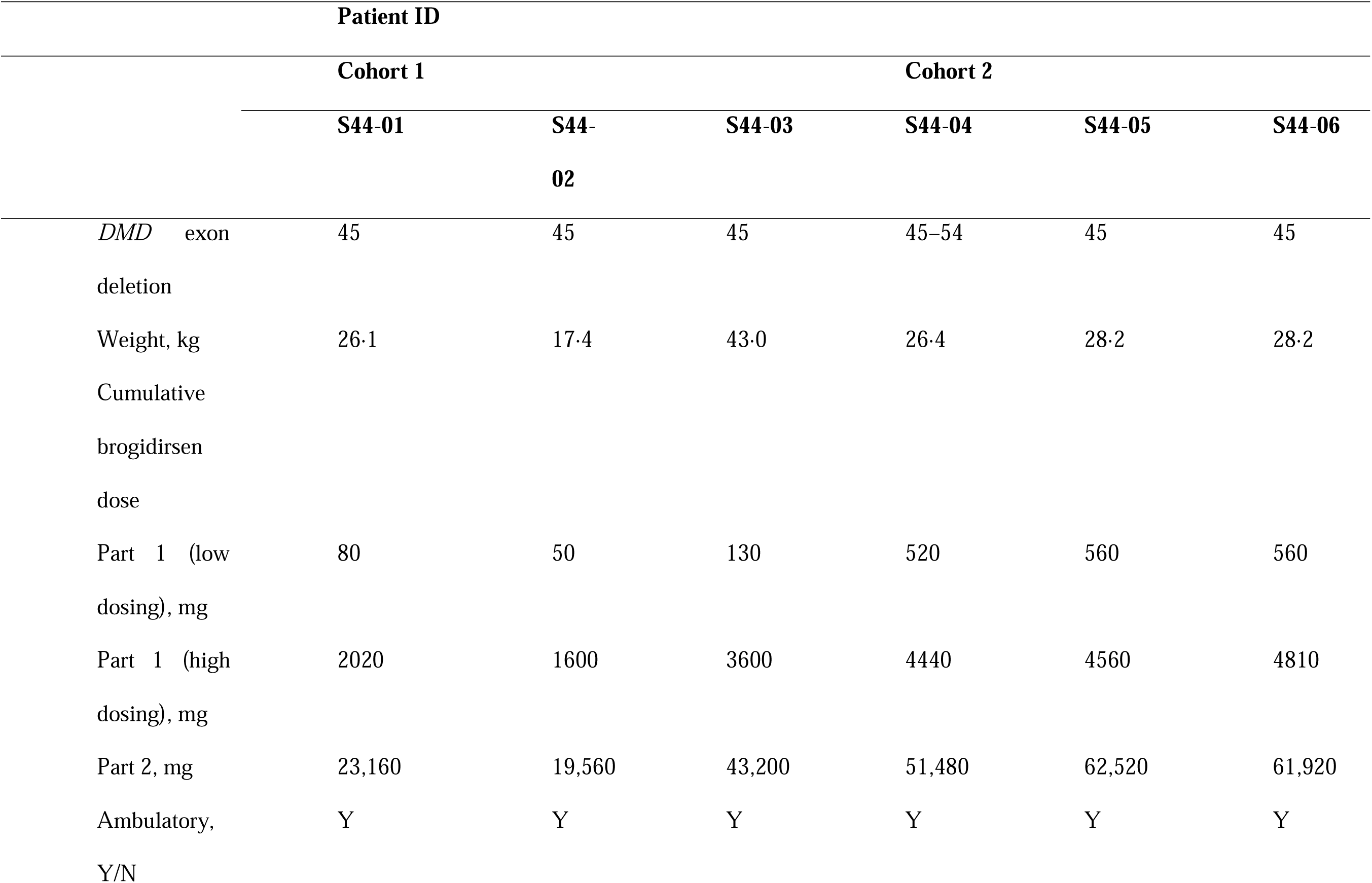
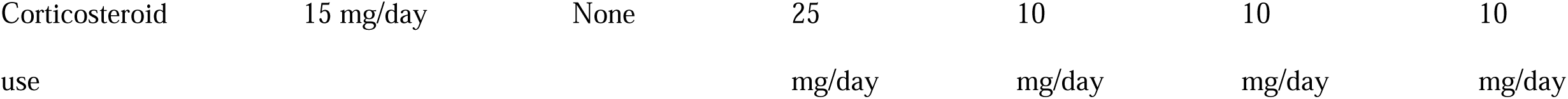
Patient background demographic and clinical characteristics and cumulative brogidirsen dose during treatment.

### Safety (primary endpoint)

The most common drug-related adverse events (AEs) were increased urinary β2 microglobulin levels, which occurred in half of the patients (all cases were in Cohort 1), and increased urine albumin/creatinine ratio and cystatin C levels, which occurred in two patients each (all cases were in Cohort 1) (**Table 2**). Only one drug-related AE was reported with the highest dose of brogidirsen (80 mg/kg). There were no serious AEs (SAEs) related to the study drug during the study and no safety or tolerability concerns related to the 40 or 80 mg/kg doses over the 24-week dose-finding stage.

**Table 2:**
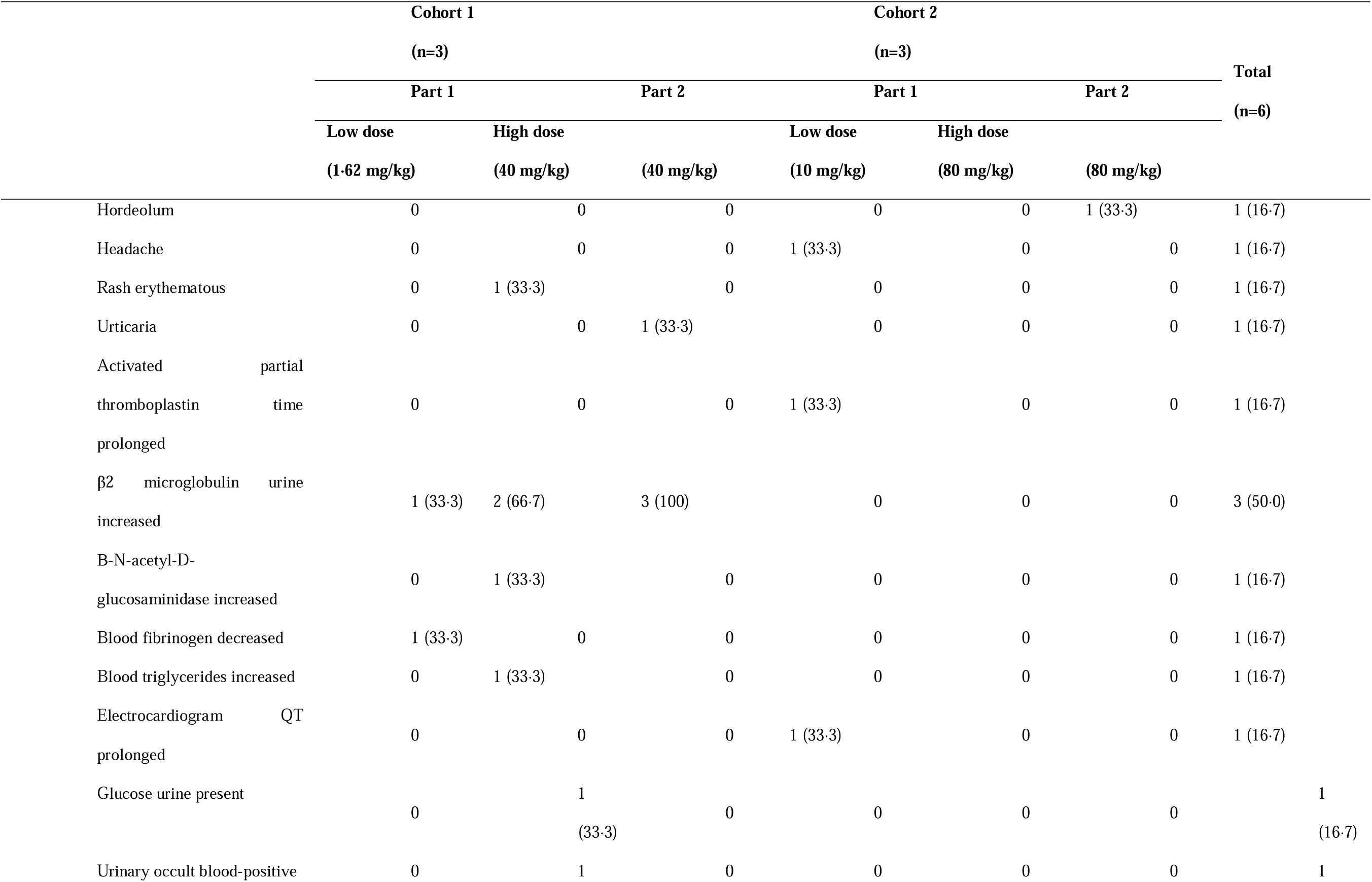
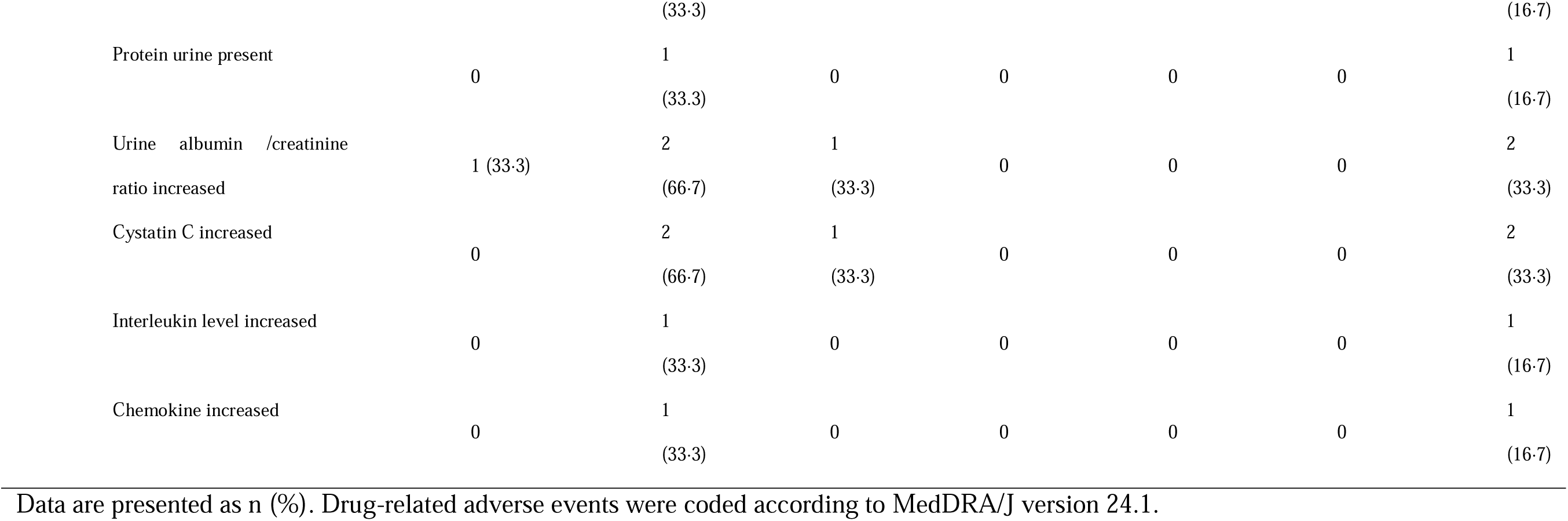
Drug-related adverse events.

|  |  | Cohort 1 |  |  | Cohort 2 |  |  | Total<br>(n=6) |
| --- | --- | --- | --- | --- | --- | --- | --- | --- |
|  |  | (n=3) |  |  | (n=3) |  |  |  |
|  |  | Part 1 |  | Part 2 | Part 1 |  | Part 2 |  |
|  |  | Low dose<br>(1·62 mg/kg) | High dose<br>(40 mg/kg) | (40 mg/kg) | Low dose<br>(10 mg/kg) | High dose<br>(80 mg/kg) | (80 mg/kg) |  |
|  | Hordeolum | 0 | 0 | 0 | 0 | 0 | 1 (33·3) | 1 (16·7) |
|  | Headache | 0 | 0 | 0 | 1 (33·3) | 0 | 0 | 1 (16·7) |
|  | Rash erythematous | 0 | 1 (33·3) | 0 | 0 | 0 | 0 | 1 (16·7) |
|  | Urticaria | 0 | 0 | 1 (33·3) | 0 | 0 | 0 | 1 (16·7) |
|  | Activated partial<br>thromboplastin time<br>prolonged | 0 | 0 | 0 | 1 (33·3) | 0 | 0 | 1 (16·7) |
|  | β2 microglobulin urine<br>increased | 1 (33·3) | 2 (66·7) | 3 (100) | 0 | 0 | 0 | 3 (50·0) |
|  | B-N-acetyl-D-glucosaminidase increased | 0 | 1 (33·3) | 0 | 0 | 0 | 0 | 1 (16·7) |
|  | Blood fibrinogen decreased | 1 (33·3) | 0 | 0 | 0 | 0 | 0 | 1 (16·7) |
|  | Blood triglycerides increased | 0 | 1 (33·3) | 0 | 0 | 0 | 0 | 1 (16·7) |
|  | Electrocardiogram QT<br>prolonged | 0 | 0 | 0 | 1 (33·3) | 0 | 0 | 1 (16·7) |
|  | Glucose urine present | 0 | 1<br>(33·3) | 0 | 0 | 0 | 0 | 1<br>(16·7) |
|  | Urinary occult blood-positive | 0 | 1 | 0 | 0 | 0 | 0 | 1 |

|  |  |  |  |  |  |  |  |
| --- | --- | --- | --- | --- | --- | --- | --- |
|  |  | (33.3) |  |  |  |  | (16.7) |
| Protein urine present | 0 | 1<br>(33.3) | 0 | 0 | 0 | 0 | 1<br>(16.7) |
| Urine albumin /creatinine<br>ratio increased | 1 (33.3) | 2<br>(66.7) | 1<br>(33.3) | 0 | 0 | 0 | 2<br>(33.3) |
| Cystatin C increased | 0 | 2<br>(66.7) | 1<br>(33.3) | 0 | 0 | 0 | 2<br>(33.3) |
| Interleukin level increased | 0 | 1<br>(33.3) | 0 | 0 | 0 | 0 | 1<br>(16.7) |
| Chemokine increased | 0 | 1<br>(33.3) | 0 | 0 | 0 | 0 | 1<br>(16.7) |

Data are presented as n (%). Drug-related adverse events were coded according to MedDRA/J version 24.1.

### Secondary endpoints

The exon 44 skipping efficiency in the biopsied muscles increased from baseline to post-treatment in all six patients (**Fig. 1A**). While the mean change from baseline was not significant for either cohort in Part 2, the mean change from baseline was substantial for the total population (mean [standard deviation (SD)], 32.10% [8.71%]; p=0.03). Additionally, the mean change from baseline was higher in Cohort 1 (80 mg/kg) than in Cohort 2 (40 mg/kg) (mean [SD], 34.64% [9.65%] vs. 29.56% [8.77%]).

**Fig. 1.**
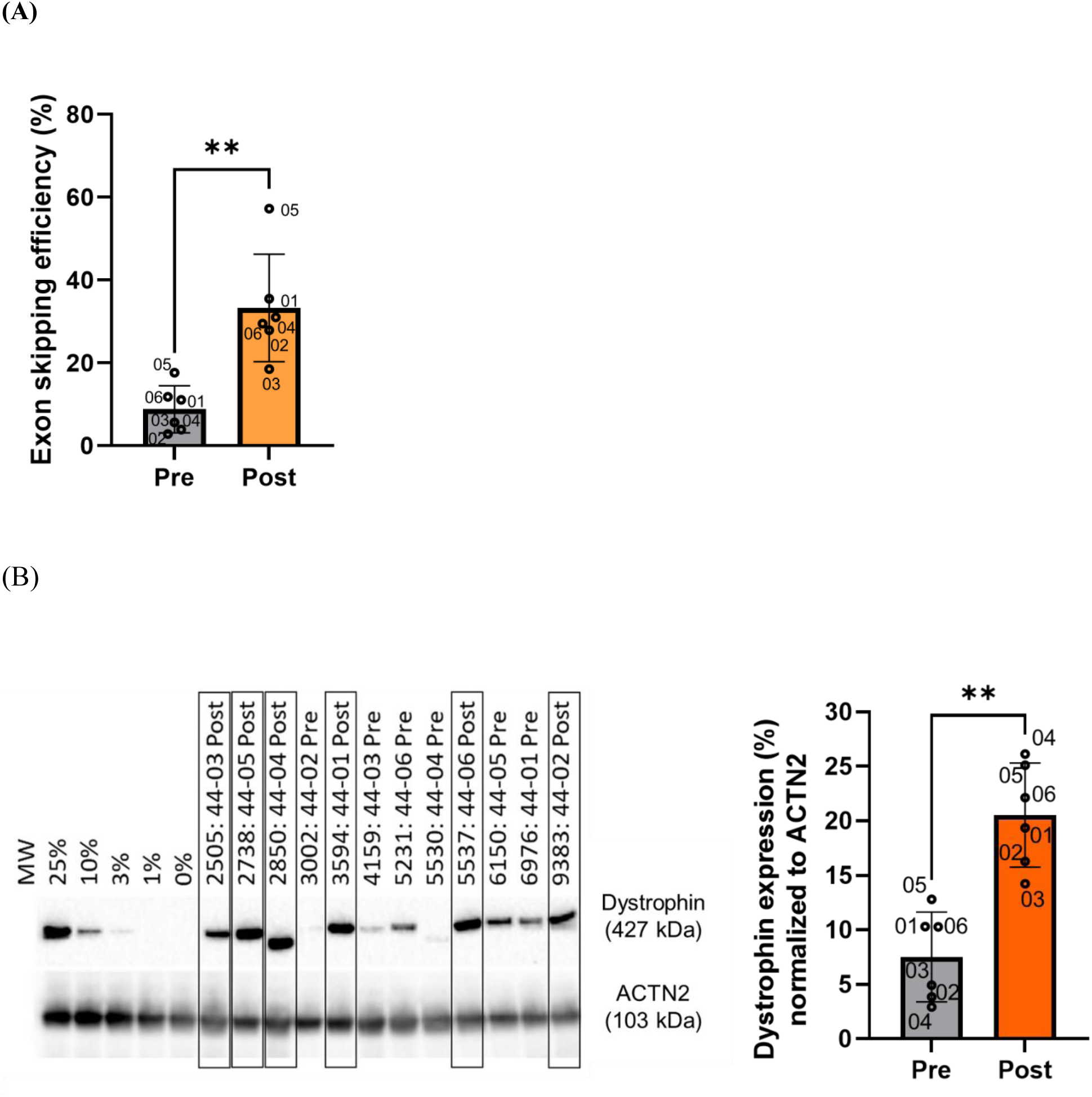
Evaluation of pre and post-treatment (24 weeks) (A) exon 44 skipping and (B) dystrophin protein expression in muscle biopsy tissue. Muscle biopsy tissues were collected from the biceps brachii of each patient before brogidirsen treatment (pre) and after completing 24 weeks of treatment (post). Exon 44 skipping was determined using reverse transcription polymerase chain reaction assays and Dystrophin protein expression was evaluated using western blotting. The size of dystrophin observed on the western blot varies because of alternative splicing. Mann-Whitney test was used for statistical analysis. Data expressed as mean ± SD; **P < 0.01. MW = molecular weight marker. SD=standard deviation. RT-PCR = reverse transcription polymerase chain reaction.

The dystrophin protein expression level in the biopsied muscles increased compared to baseline with treatment in each patient, and the increase was significant in the total population (mean [SD], 13.03% [5.21%]; p=0.03) (**Fig. 1B**). Furthermore, there was a noteworthy trend for a higher change in dystrophin expression in Cohort 2 than in Cohort 1 (mean [SD], 15.79% [6.44%] vs. 10.27% [1.88%]).

The serum creatinine kinase levels, a biomarker of muscle damage or degeneration, tended to decrease over time with treatment (**Fig. S1**). No apparent improvement in motor function was observed with treatment; however, the patients maintained their motor function and showed a general trend towards improvement (**Fig. 2**). Moreover, there was an apparent dose-dependent increase in C_max_ and AUC_0–t_ and no evident accumulation of brogidirsen over time (**Table 3**).

**Fig. 2.**
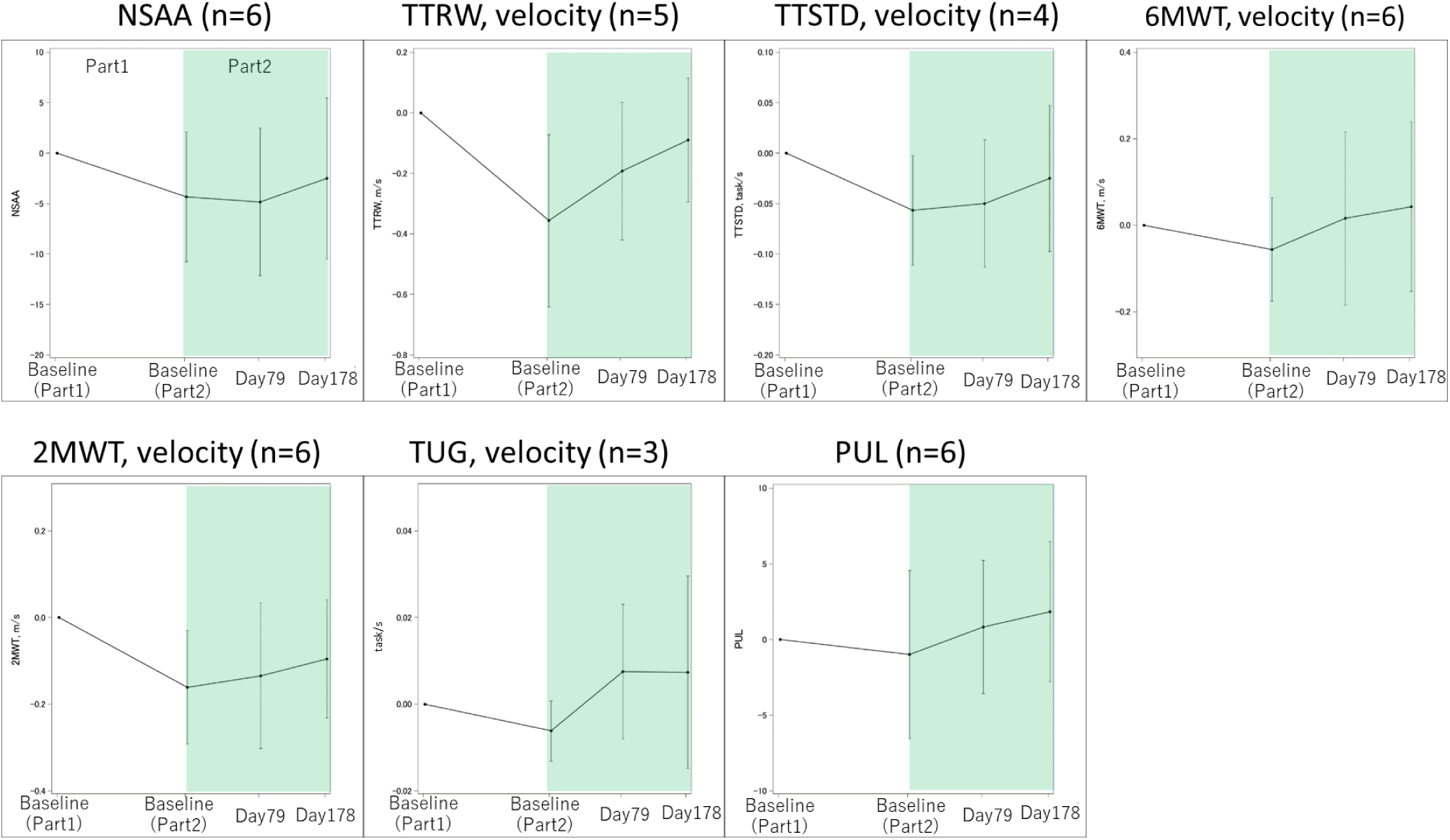
Changes from baseline in motor function tests. Increasing the score in all the tests represented an improvement in motor functions. 2MWT = 2-minute walk test. 6MWT = 6-minute walk test. NSAA = North Star Ambulatory Assessment score. PUL = Performance of Upper Limb. TTRW = time to run/walk 10 meters. TTSTD = time to stand from the supine test. TUG = timed “up & go” test.

**Table 3:** Brogidirsen pharmacokinetics.

|  | Part 1, initial dose |  |  |  | Part 2, initial dose |  | Part 2, 24<sup>th</sup> dose |  |
| --- | --- | --- | --- | --- | --- | --- | --- | --- |
|  | Cohort 1 |  | Cohort 2 |  | Cohort 1 | Cohort 2 | Cohort 1 | Cohort 2 |
|  | (n=3) |  | (n=3) |  | (n=3) | (n=3) | (n=3) | (n=3) |
|  | 1·62 mg/kg | 40 mg/kg | 10 mg/kg | 80 mg/kg | 40 mg/kg | 80 mg/kg | 40 mg/kg | 80 mg/kg |
| C<sub>max</sub> (µg/mL) | 4·85 ± 0·79 | 141 ± 24 | 32·9 ± 3·6 | 316 ± 59 | 170 ± 14 | 273 ± 71 | 168 ± 10 | 299 ± 113 |
| T<sub>max</sub> (h) | 1·00 ± 0 | 1·00 ± 0 | 1·00 ± 0 | 1·00 ± 0 | 1·00 ± 0 | 0·833 ± 0·289 | 1·00 ± 0 | 0·677 ± 0·289 |
| T<sub>1/2</sub> (h) | 1·65 ± 0·11 | 2·78 ± 0·13 | 2·00 ± 0·89 | 2·67 ± 0·32 | 2·77 ± 0·16 | 2·69 ± 0·18 | 2·70 ± 0·11 | 2·84 ± 0·22 |
| AUC<sub>0-t</sub> (µg·h/mL) | 7·63 ± 1·54 | 231 ± 34 | 55·1 ± 13·6 | 503 ± 155 | 222 ± 18 | 497 ± 223 | 244 ± 30 | 519 ± 239 |
| CL (mL/h/kg) | 216 ± 44 | 176 ± 28 | 187 ± 41 | 169 ± 51 | 181 ± 15 | 182 ± 73 | 166 ± 20 | 174 ± 65 |
| Vz (mL/kg) | 508 ± 82 | 704 ± 116 | 507 ± 98 | 643 ± 179 | 722 ± 74 | 697 ± 238 | 644 ± 83 | 702 ± 243 |

Data are presented as mean ± standard deviation.
AUC<sub>0-t</sub>= area under the plasma concentration-time curve from the start of administration to the last quantifiable plasma concentration point.
CL=drug clearance. C<sub>max</sub>=maximum plasma concentration. t<sub>1/2</sub>=half-life. T<sub>max</sub>=time to reach C<sub>max</sub>. Vz = volume of distribution based on the terminal phase.

### High-throughput proteomic assays of plasma

Plasma collected from six patients in Part 2 of the study was analysed by the Olink® Explore 3072 high-throughput proteomics platform at three different time points: baseline and 12 and 24 weeks. Three proteins were upregulated, and 11 were downregulated at week 12 compared to baseline (**Fig. 3A**). Moreover, two proteins were upregulated and 55 were downregulated among 2944 proteins at week 24 compared to baseline (**Fig. 3B**). The levels of plasma markers titin (TTN), myomesin 2 (MYOM2), and myosin light chain, phosphorylatable, fast skeletal muscle (MYLPF), which were elevated in patients with DMD,^39,40^ decreased in Part 2 in all patients (**Fig. 3C, 3D, 3E**, and **3F**). The levels of dystrophin or inflammatory cytokines, including interleukin 6 (IL6), tumour necrosis factor (TNF), interferon-gamma (IFNG), matrix metallopeptidase 9 (MMP9), and osteopontin (secreted phosphoprotein 1, SPP1), showed slight changes in Part 2 in all patients, except for one (44-02) who did not receive steroid treatment (Supplementary Appendix, **Fig. S2A, S2B, S2C, S2D, S2E, and S2F**). Ciliary neurotrophic factor (CNTF), a homolog of IL6, was upregulated, and the calcium-dependent enzyme, Peptidyl arginine deiminase 2 (PADI2), was downregulated after treatment (**Fig. 3G** and **3H**).

**Fig. 3.**
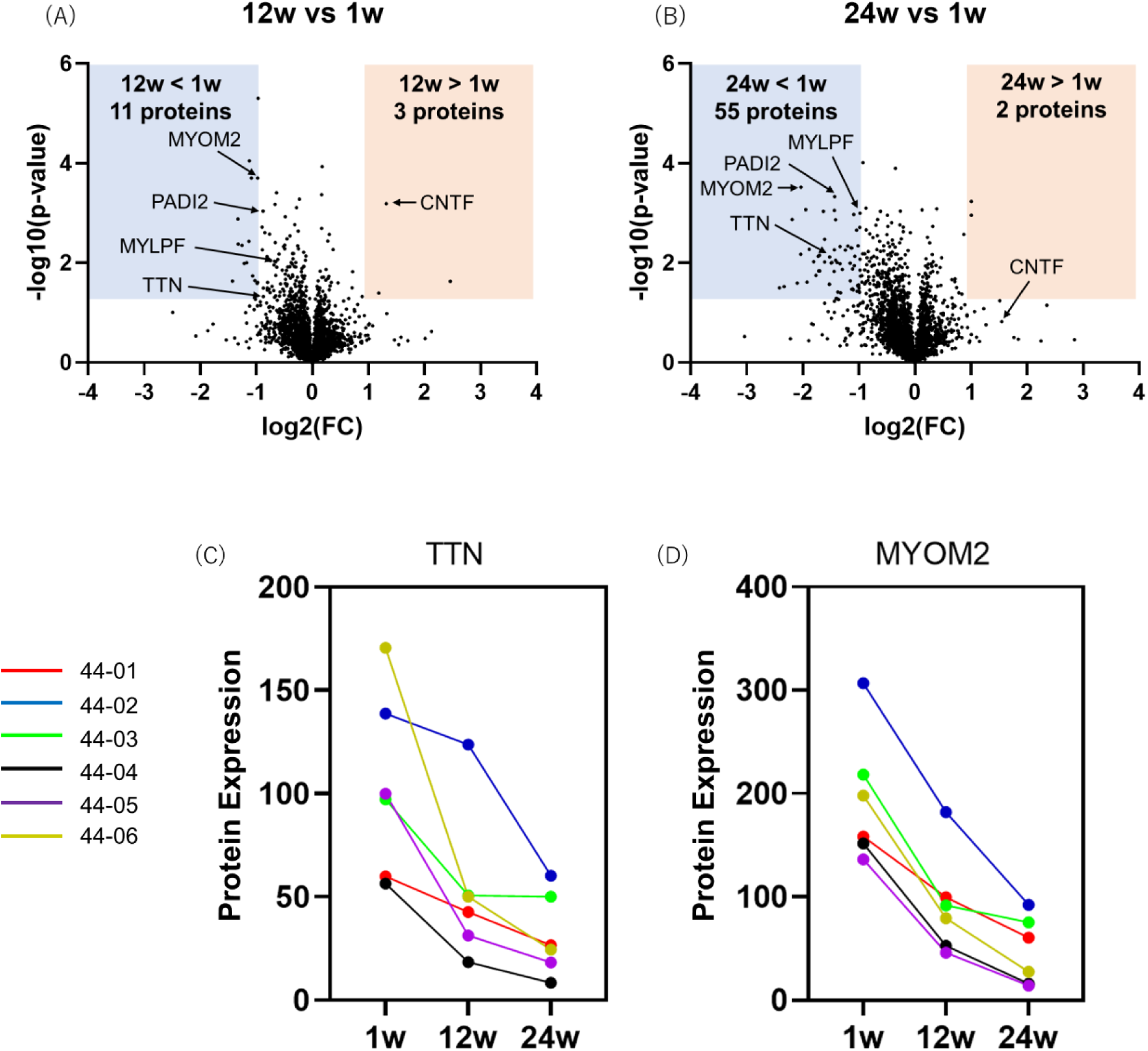
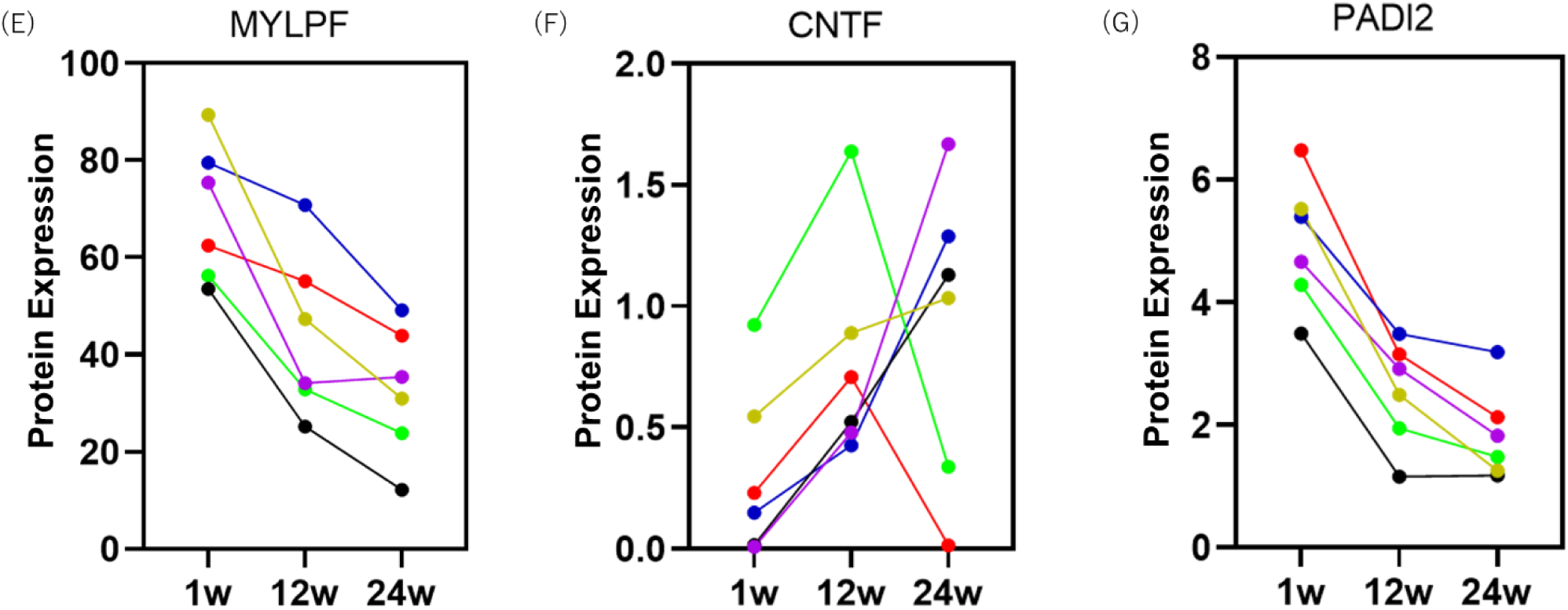
Changes of proteomics in plasma during treatment (A) volcano plots (12 weeks versus one week), (B) volcano plots (24 weeks versus one week), temporal profile of (C) TTN, (D) MYOM2, (E) MYLPF, (F) CNTF and (G) PADI2. Plasma was collected from each patient at three different time points, including 1 week, 12 weeks, and 24 weeks in Part 2. Protein expression was evaluated using proximity extension assays (Olink) and shown as 2^NPX^. NPX = normalised protein expression.

### In vitro assay

*In vitro* testing of brogidirsen treatment for three types of patient-derived cells, including MYOD1-UDCs, myotubes and MYOD1-transduced fibroblasts, was conducted. The results demonstrated that the exon skipping efficiency and dystrophin protein expression increased dose-dependent in all cell types from six patients (**Fig. 4A** and **4B**; Supplementary Appendix, **Fig. S3A** and **S3B**). Alternative skipping of exon 44 and dystrophin protein expression were observed before brogidirsen treatment. The half-maximal effective concentration (EC_50_) (skipping efficiency) of brogidirsen was 7.12 µM (**Fig. 4C**).

**Fig. 4.**
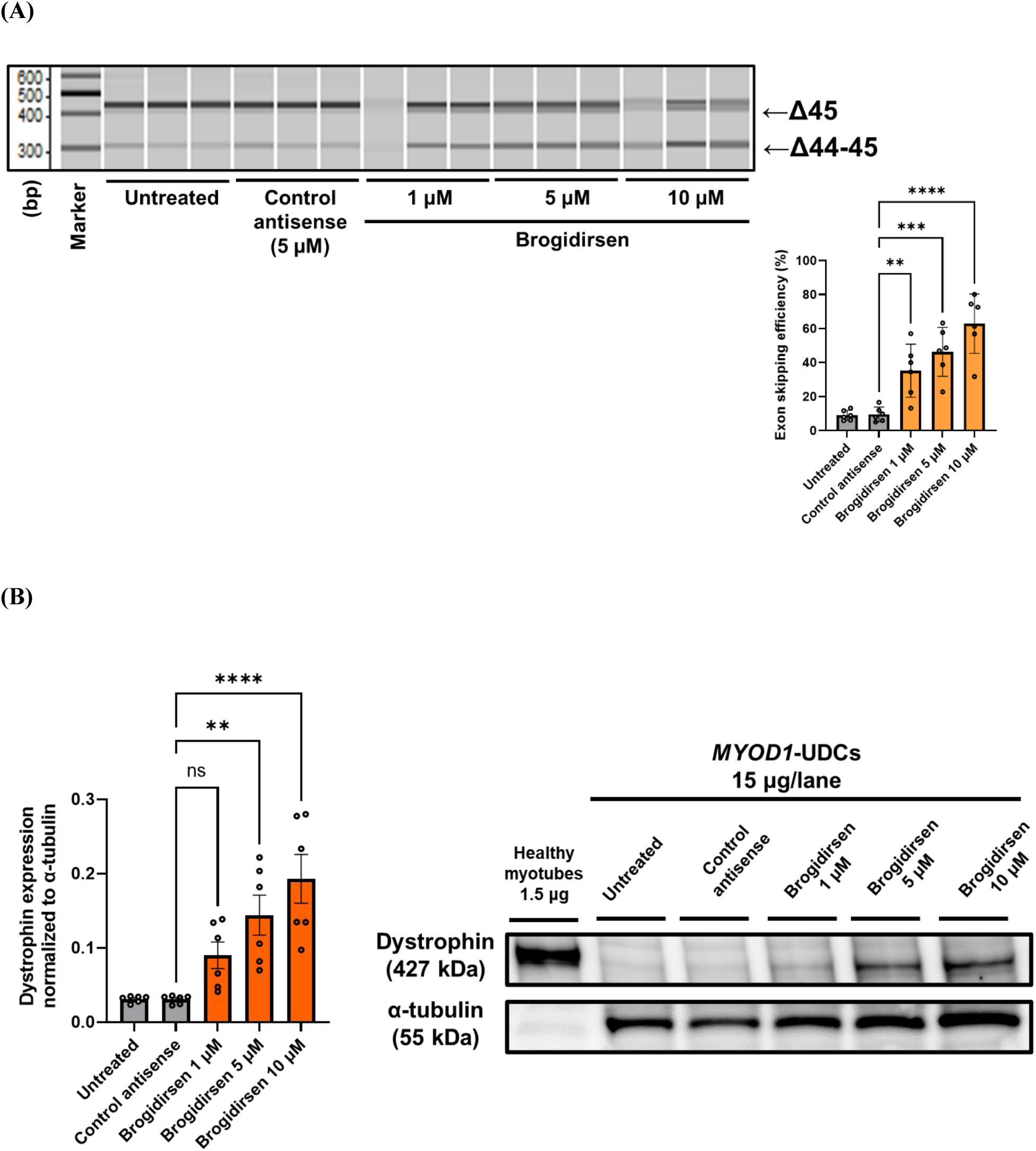
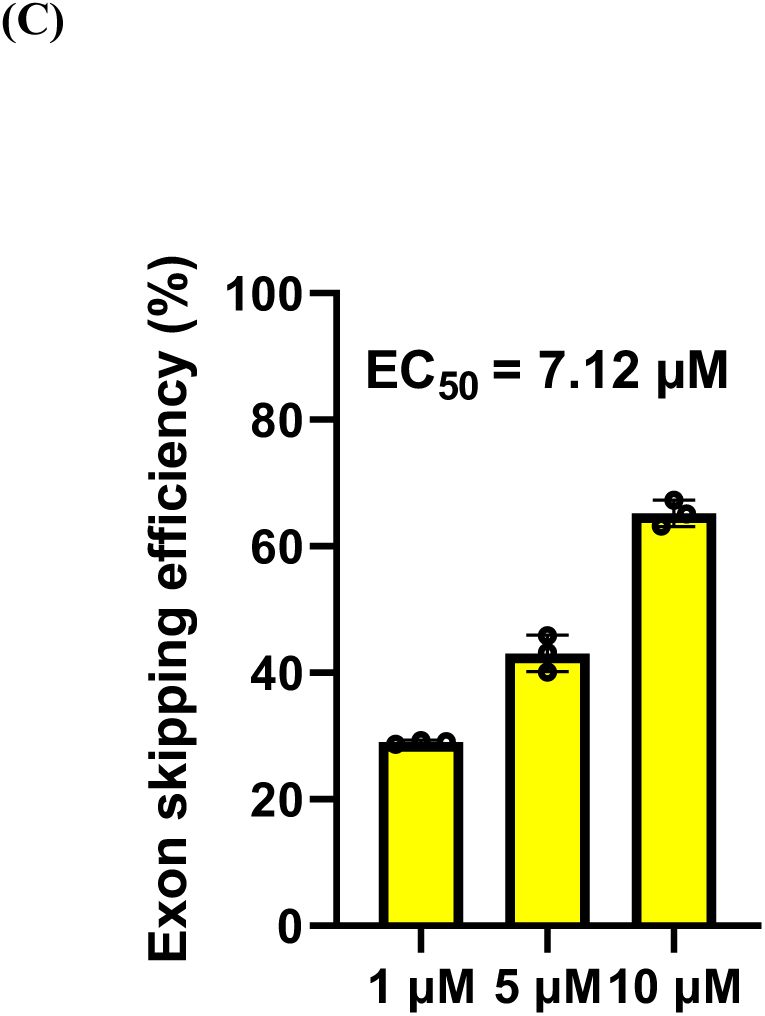
Evaluation of (A) exon 44 skipping in patient-derived MYOD1-UDCs. (B) dystrophin expression in patient-derived MYOD1-UDCs following *in vitro* treatment with brogidirsen. (C) Determination of EC_50_ of brogidirsen in healthy donor-derived MYOD1-UDCs. Exon 44 skipping was determined using reverse transcription polymerase chain reaction assays, and dystrophin protein expression was evaluated using western blotting. Significant differences between treatment with brogidirsen and control antisense PMO were determined using Dunnett’s multiple comparisons test. Data expressed as mean ± SD; **P < 0.01, ***P<0.001 and ****P<0.0001. EC_50_ = half maximal effective concentration. PMO = phosphorodiamidate morpholino oligonucleotide. SD = standard deviation. UDC = urine-derived cell.

## DISCUSSION

This first-in-human study evaluating brogidirsen in patients with DMD demonstrated that brogidirsen was safe and well tolerated at doses of up to 80 mg/kg for 24 weeks. No SAEs were reported, and none of the patients discontinued the study. With treatment, exon 44 skipping efficiency and dystrophin protein expression were higher than the baseline.

The dystrophin expression level in the present study was higher than in previous studies, wherein patients with DMD were subjected to exon-skipping therapy. Dystrophin levels were 16.63% and 24.47% higher than the normal levels after 24-week treatment with brogidirsen at 40 mg/kg and 80 mg/kg, respectively. The respective levels of dystrophin protein recovery after treatment were 10.27% and 15.79%. These levels exceeded those reported in a phase II study involving patients with DMD treated with viltolarsen for 20–24 weeks (40 mg/kg group, 5.7%; 80 mg/kg group, 5.9%).^41,42^

Hoffman et al. proposed a classification system for patients with suspected Becker muscular dystrophy (BMD) that categorised patients as having mild BMD if the dystrophin protein level was >20%.^43^ Considering that patients treated with 80 mg/kg brogidirsen in our study had dystrophin expression levels >20% of the normal levels after 24 weeks of treatment, we expect an excellent functional prognosis with this treatment. Furthermore, a report of families with X-linked dilated cardiomyopathy with a DMD mutation suggested that skeletal muscle symptoms could be avoided with a dystrophin expression level of ≥30% of normal levels.^44^ Moreover, a long-term study on exon-skipping therapy reported that dystrophin expression might accumulate and increase with long-term repeated administration of golodirsen.^45^ Importantly, no anti-dystrophin antibodies were detected (data not shown), and serum creatinine kinase levels tended to decrease with drug administration.

In Part 1 of the present study, where relatively low doses of brogidirsen were administered over 4 weeks, there was a trend towards decreased motor function. However, in Part 2, when the dose was higher and continued for 24 weeks, there was a trend towards toward maintaining or slightly improving motor function. Considering that all the included patients could walk before treatment, caution is needed when interpreting the efficacy of brogidirsen in terms of motor function. Although DMD progresses relatively slowly in patients with exon 44 skipping, a natural history study conducted by Brogna et al. suggests that motor function declines when patients with DMD, including those with exon 44 skipping, are ≥7 years of age.^46^ Considering that five of the six patients in the present study were ≥8 years old, brogidirsen may have been effective simply because the motor function test scores were maintained or slightly improved from baseline through completing 24 consecutive weeks of treatment in Part 2. Thus, while there were no significant differences in the changes in motor function scores from baseline to post-treatment, the authors consider these findings to be clinically significant, given the small number of patients in the study (N=6) and the short duration of treatment.

The results of the first-in-human study of viltolarsen suggested that there were both responders and non-responders to treatment, as indicated by the degree of exon skipping and dystrophin protein recovery.^41^ None of the patients in the present study were non-responders. The EC_50_ value of brogidirsen was obtained using MYOD1-UDCs from healthy volunteers and revealed that brogidirsen was highly active, supporting the absence of non-responders in the present study. Before initiating this study, we were interested in understanding whether the *in vitro* efficiency of exon skipping and dystrophin recovery following brogidirsen treatment could predict the effects of the drug on human muscle tissues *in vivo*. Given that there were no non-responders, we could only discuss efficacy among the responders. However, this was challenging due to the difference in doses between Cohorts 1 and 2, and only one patient had a deletion of DMD exons 45–54.

Additionally, DMD exon 45 and 45–54-deletion patients showed naturally occurring exon 44 skipping due to alternative splicing and dystrophin protein expression before brogidirsen administration. It is known that DMD patients amenable to exon 44 skipping had an elevated number of revertant and trace dystrophin expression (approximately 19% of control, using quantitative immunohistochemistry) presenting with intermediate muscular dystrophy or a BMD-like phenotype.^47^ This differed from what was observed in patients with deletions in DMD exons 48–52, 45–52, and 49–52 in the first-in-human study involving viltolarsen.^41^ These differences in naturally occurring exon 44 skipping and dystrophin expression between the two studies may have influenced our results.

We identified several candidates’ plasma markers responsive to exon-skipping therapy using the Olink® Explore 3072 high-throughput proteomic analysis of plasma. Among the small number of proteins that exhibited significant changes during treatment, the levels of TTN, MYOM2, and MYLPF decreased. The plasma markers are associated with muscle tissue necrosis,^39,40^ and a decrease in their levels may indicate an improvement in muscle pathology. While we expected a reduction in the levels of the inflammatory markers, including IL6, TNF, IFNG, MMP9, and SPP1, which are typically elevated in the blood of patients with DMD,^48,49,50^ we did not observe significant changes in Part 2 of our study. All patients, except 44-02, were treated with steroids; therefore, inflammation might have been alleviated. However, our data showed that the levels of plasma CNTF, the IL6 homolog, were elevated, and motor function was maintained in Part 2. Low serum CNTF levels are associated with severe clinical phenotypes in patients with DMD without steroid treatment.^51^ CNTF may help monitor follow-up responsiveness to exon skipping, unlike other inflammatory markers, even in steroid-treated patients. Furthermore, the level of PADI2, which is elevated in DMD astrocytes,^52^ was significantly reduced in Part 2 in all the patients. The biological significance of PADI2 changes in DMD remains unclear; however, we can utilise this as a novel therapeutic marker in trials for DMD.

This is the first study to use MYOD1-UDCs from study patients to evaluate the efficacy of a drug in a DMD clinical trial. The results of the *in vitro* assays using MYOD1-UDCs suggest that exon 44 skipping and the recovery of dystrophin protein levels can be achieved when an adequate concentration of brogidirsen reaches the skeletal muscle, consistent with its *in vivo* efficacy. These findings confirm the dose-dependent exon skipping and dystrophin protein recovery of brogidirsen in MYOD1-UDCs (myotubes generated from UDCs)^37^ obtained by noninvasive urine collection. This achievement is significant, as future clinical trials can utilise this methodology to evaluate the pharmacological efficacy and sequence optimisation of PMO drugs and determine patient eligibility for specific PMO treatments.

Exon-skipping therapy is limited to patients with specific genetic mutations and requires weekly intravenous administration.^53^ The treatment burden of patients with DMD may be reduced with newer formulations that improve muscle delivery and blood retention, such as conjugation with a safe cell-penetrating peptide or chemical modifications to PMO, reducing the frequency of administration. Considering that recent clinical developments in the treatment of DMD have shifted towards gene therapy and that exon-skipping technologies use new-generation chemical compounds, the promising balance between safety and efficacy observed in the present study suggests that conventional PMO-based exon-skipping treatments should receive renewed attention. These findings may guide a change in future clinical developments in treating DMD.

The limitations include the small number of patients, short treatment period, and open-label study design. Although none were observed in the present study, future studies on brogidirsen should also monitor for non-responders. Additionally, future studies are required to further evaluate whether the efficacy of brogidirsen can be predicted *in vitro* using MYOD1-UDCs from patients who are potential treatment candidates.

In conclusion, this first-in-human studies of brogidirsen in six ambulant patients with DMD amenable to exon 44 skipping demonstrated that brogidirsen was safe and had favourable pharmacokinetics. All six patients observed notable improvements in exon 44 skipping and dystrophin protein expression after treatment. An extension of this study is currently underway (NCT05135663).

## Data Availability

All data produced in the present study are available upon reasonable request to the authors.

## Acknowledgements

We thank the study participants and their families for participating in the trial. The authors thank Sarah Bubeck, PhD, of Edanz (www.edanz.com) for providing medical writing support, which was funded by grants from the Japan Agency for Medical Research and Development (AMED) awarded to Y.A. (18lm0203066h0001, 21lm0203086h0003), in accordance with Good Publication Practice (GPP3) guidelines (http://www.ismpp.org/gpp3). We thank North Star Clinical Network for the North Star Ambulatory Assessment score and Performance of Upper Limb evaluations. We thank AGADA Bioscience Inc. for the western blot analysis of muscle tissues. We thank Shin Nippon Biomedical Laboratories, Ltd. for exon skipping analysis of muscle tissues. The National Center of Neurology and Psychiatry (NCNP) and Nippon Shinyaku Co., Ltd. are the co-inventors of NS-089/NCNP-02, brogidirsen. The phase I/II study was funded by a grant from the Japan Agency for Medical Research and Development (AMED) awarded to Y.A. (grant number 21lm0203086h0003). The phase II extension study is being funded by Nippon Shinyaku Co., Ltd.

## Author contributions

Conceptualisation: K.K., Y.A.

Methodology: K.K., S.M., E.H

Investigation: H.K., E.T., T.I., Y.S-M., A.I., M.S., C.Y.

Visualisation: K.K., S.M., E.H

Funding acquisition: Y.A.

Project administration: Y.A.

Supervision: Y.A.

Writing – original draft: K.K., Y.A.

Writing – review & editing: H.K., E.T., T.I., Y.S-M., A.I., M.S., C.Y., S.M., E.H

## STAR METHODS

### Study design and treatments

The study protocol for this open-label, dose-escalation, two-centre, phase I/II trial is currently available.^54^ The study was conducted in two parts **(Fig. S4)**. Part 1 was conducted over 2 weeks with stepwise dose escalation that included four doses of brogidirsen (dose level 1, 1.62 mg/kg once weekly [QW]; dose level 2, 10 mg/kg QW; dose level 3, 40 mg/kg QW; and dose level 4, 80 mg/kg QW). Part 2 was conducted over 24 weeks to evaluate the determined dose. Brogidirsen was administered intravenously over 1 h (±10 min).

Part 1 included two cohorts: Cohort 1 received the study drug at 1·62 mg/kg QW (dose level 1) for 2 weeks, followed by a dose increase to 40 mg/kg QW (dose level 3) for 2 weeks; and Cohort 2 received the study drug at 10 mg/kg QW (dose level 2) for the first 2 weeks and 80 mg/kg QW (dose level 4) for the next 2 weeks. Three patients from each cohort were evaluated. The two doses used in Part 2 were determined based on the findings from Part 1. In Part 2, brogidirsen was administered QW intravenously for 24 weeks to three patients in each cohort (the dose group). In an ongoing extension study, the patients will receive 40 or 80 mg/kg intravenously for 72 weeks.

The patients continued physiotherapy or exercise therapy regimens (unchanged) from 90 days before treatment initiation through the end of the post-treatment observation period. Pre-existing concomitant therapies, including steroids, were allowed from enrollment until the end of the observation period. All other investigational drugs, idebenone, coenzyme Q10 products, and adenosine triphosphate products were prohibited. The study included a pre-treatment period of up to 13 weeks, a treatment period, and a post-treatment period of 6 weeks.

This study complied with the ethical principles of the Declaration of Helsinki and all applicable Japanese local and national regulatory laws. The study protocol and related documentation were approved by the Institutional Review Board of the NCNP (approval number II-012). The study was registered at clinicaltrials.gov (NCT04129294) and the University Hospital Medical Information Network (UMIN000038505). Brogidirsen was synthesised and purified by Nippon Shinyaku Co., Ltd. It was supplied at 50 mg/mL in 3 mL vials. The other published article provided detailed information on the drug, including the sequences.^36^ Nippon Shinyaku Co., Ltd. had no role in collecting, analysing, interpreting data, writing the report, or submitting the paper for publication.

### Patients

The key inclusion criteria were male sex, 4 to <17 years old, life expectancy ≥1 year, ability to undergo muscle biopsy from biceps brachii or tibialis anterior, presence of a multiplex ligation-dependent probe amplification (MLPA)-confirmed out-of-frame deletion amenable to exon 44 skipping of the DMD gene, a corrected QT interval (QTc) <450 ms (standard 12-lead electrocardiogram) or QTc <480 ms for patients with bundle branch block, and if the patient used systemic corticosteroid therapy, it must have been started ≥6 months prior to study enrollment with no dosage changes within the 3 months prior to enrollment. The key exclusion criteria were <50% of the predicted value (based on the reference values for spirometry in Japanese children)^55^ for forced vital capacity and <40% left ventricular ejection fraction, or <25% fractional shortening (by echocardiogram). The full list of the exclusion criteria is available in the Supplementary Appendix, **Text S1**. The legal representatives of each patient provided written informed consent prior to study participation, and patients provided voluntary assent wherever possible.

### Outcomes

The primary (safety) endpoints were findings from physical examinations, vital signs, 12-lead electrocardiography, echocardiography, and adverse event reporting. Measurements included the DMD gene test (MLPA analysis), exon 44 sequencing analysis, haematology, blood chemistry, urinalysis, cytokine/complement levels, immunology tests, and pulmonary function tests. The Medical Dictionary for Regulatory Activities version 23.1 was used to code the AEs.

The secondary endpoints included the efficiency of exon 44 skipping, dystrophin protein expression analysis, changes in blood creatine kinase levels, assessment of motor function (including the NSAA score, 10-meter walk/run test, time to stand from supine test, 6-minute walk test, 2-minute walk test, timed “up & go” test, and Performance of Upper Limb),^54,56^ and plasma brogidirsen concentration determination. The efficiency of exon 44 skipping was evaluated by reverse transcription polymerase chain reaction (RT-PCR) using total RNA from sliced frozen muscle. Only two primer-specific bands corresponding to fragments with and without exon 44 were selected. The exon-skipping level (%) was analysed using the Agilent 2100 Bioanalyzer Electrophoresis System (Agilent Technologies) and calculated for each primer pair as exon 44–deleted PCR fragment/ (full-length PCR fragment + exon 44–deleted PCR fragment) × 100%. The primer sequences used for each patient are shown in **Table S1**. Western blotting analysis was applied to confirm dystrophin expression using primary anti-dystrophin (1:300; Abcam, ab15277) and anti-ACTN2 (1:10000; Proteintech, 14221-1-AP) antibodies; dystrophin expression was quantified as the dystrophin/ACTN2 band intensity ratio.

Blood was collected for plasma pharmacokinetic evaluation, and the brogidirsen concentration and pharmacokinetic parameters were assessed using high-performance liquid chromatography with tandem mass spectrometry.

*In vitro* assays were conducted using cells derived from the urine, skin, and muscle tissue collected from each patient prior to treatment initiation and differentiated into myotubes.^37^ Differentiated cells were treated with brogidirsen (1, 5, or 10 µM) or a control antisense PMO. The efficiency of exon 44 skipping was evaluated by RT-PCR using total RNA from each cell type. Only two primer-specific bands corresponding to fragments with and without exon 44 were selected. The exon-skipping level (%) was analysed using the MultiNA, a microchip electrophoresis system (Shimadzu) and calculated for each primer pair as exon 44–deleted PCR fragment/ (full-length PCR fragment + exon 44–deleted PCR fragment) × 100%. The primer sequences used for each patient are shown in **Table S1**. Western blotting analysis was applied to confirm dystrophin expression using primary anti-dystrophin (1:500; Abcam, ab15277) and anti-α -tubulin (1:1200; Sigma, T6199) antibodies; dystrophin expression was quantified as the dystrophin/α-tubulin band intensity ratio.

Plasma collected from the six patients in Part 2 of the study was analysed at three-time points (baseline, 12 weeks, and 24 weeks) using the Olink Explore 3072 platform for high-throughput protein biomarker discovery: the analysis utilised proximity extension assays and quantitative polymerase chain reaction (qPCR). Pairs of antibodies with complementary oligonucleotide strands bound the target protein, and when in proximity, the strands hybridised, creating a DNA chain serving as an id tag, which was elongated with DNA polymerase and read by qPCR. Data are described as normalised protein expression (NPX; arbitrary units) on a log2 scale. Quality control (QC) was performed according to Olink’s standard protocol. Internal controls were added to each sample for the plate and sample QC. All plates and samples passed QC thresholds, an internal control SD of <0.2 NPX from the median between plates and <0.3 NPX between samples within plates.

### Statistical Analysis

Statistical significance was not considered when determining the sample size; instead, it was determined based on the feasibility of enrollment and assessment of motor function at a single centre. The safety analysis set included all patients treated at least once with brogidirsen, and the efficacy and pharmacokinetic analysis sets included all enrolled patients without serious eligibility violations and not lacking the applicable data.

Summary statistics were used to report the patients’ demographic data. For the primary analysis (safety evaluation), the number and frequency of drug-related AEs and SAEs were documented. Pharmacokinetic studies included the determination of the maximum plasma concentration (C_max_), the area under the plasma concentration-time curve from the start of administration to the last quantifiable plasma concentration point (AUC_0–t_), half-life, systemic clearance, and distribution volume for brogidirsen. The data regarding the Brogidirsen administration was documented. The Mann-Whitney test was used to determine significant differences in exon skipping efficiency and dystrophin protein expression of the biopsied muscles between treatments with brogidirsen (**Fig 2A and 2B**). Significant differences between treatments with brogidirsen and control antisense were determined using Dunnett’s multiple comparisons tests (**Fig 4A, 4B, S3A and S3B)**. Throughout this article, we considered the P value < 0.05 as statistically significant. All analyses used GraphPad Prism Version 9.2.0 (GraphPad Software, San Diego, CA, USA), SAS software version 9.4 (SAS Institute Inc., Cary, NC, USA) and Microsoft Excel 2016/2019.

---

**Figure S1: Changes in plasma creatinine kinase throughout brogidirsen treatment during Part 2.** Cohort 1 was treated with 40 mg/kg brogidirsen, and Cohort 2 was treated with 80 mg/kg brogidirsen (administered intravenously once weekly for 24 weeks). CK=creatinine kinase.

**Figure S2: Changes of proteomics in plasma during treatment (A) temporal profile of Dystrophin, (B) IL6, (C) TNF, (D) IFNG, (E) SPP1 (osteopontin) and (F) MMP9.** Plasma was collected from each patient at 3 different time points, including 1 week, 12 weeks, and 24 weeks in Part 2. Protein expression was evaluated using proximity extension assays (Olink).

**Figure S3: Evaluation of (A) exon 44 skipping and dystrophin protein expression in patient-derived myoblast-derived myotubes and (B) patient-derived MYOD1 fibroblasts following *in vitro* treatment with brogidirsen**

**Figure S4: Study design**

**Table S1: List of primers**

## References

1. E. P. Hoffman, R. H. Brown Jr, L. M. Kunkel, Dystrophin: The protein product of the Duchenne muscular dystrophy locus. Cell. 51(6), 919–928 (1987).

2. Q. Q. Gao, E. M. McNally, The Dystrophin Complex: Structure, Function, and Implications for Therapy. Compr. Physiol. 5(3), 1223–1239 (2015).

3. E. Mercuri, C. G. Bönnemann, F. Muntoni, Muscular dystrophies. Lancet. 394(10213), 2025–2038 (2019).

4. S. Crisafulli, J. Sultana, A. Fontana, F. Salvo, S. Messina, G. Trifirò, Global epidemiology of Duchenne muscular dystrophy: an updated systematic review and meta-analysis. Orphanet. J. Rare. Dis. 15(1), 141 (2020).

5. S. Ryder, R. M. Leadley, N. Armstrong, M. Westwood, S. de Kock, T. Butt, M. Jain, J. Kleijnen, The burden, epidemiology, costs and treatment for Duchenne muscular dystrophy: an evidence review. Orphanet. J. Rare. Dis. 12(1). 79 (2017).

6. D. Duan, N. Goemans, S. Takeda, E. Mercuri, A. Aartsma-Rus, Duchenne muscular dystrophy. Nat. Rev. Dis. Primers. 7(1), 13 (2021).

7. T. C. Roberts, R. Langer, M. J. A. Wood, Advances in oligonucleotide drug delivery. Nat. Rev. Drug. Discov. 19(10), 673–694 (2020).

8. S. Takeda, P. R. Clemens, E. P. Hoffman, Exon-Skipping in Duchenne Muscular Dystrophy. J. Neuromuscul. Dis. 8(s2), S343–S358 (2021).

9. A. Aartsma-Rus, I. Fokkema, J. Verschuuren, I. Ginjaar, J. van Deutekom, GJ. van Ommen, J. T. den Dunnen, Theoretic applicability of antisense-mediated exon skipping for Duchenne muscular dystrophy mutations. Hum. Mutat. 30(3), 293–299 (2009).

10. M. J. Wood, Toward an oligonucleotide therapy for Duchenne muscular dystrophy: a complex development challenge. Sci. Transl. Med. 2(25), 25ps15 (2010).

11. A. Aartsma-Rus, V. Straub, R. Hemmings, M. Haas, G. Schlosser-Weber, V. Stoyanova-Beninska, E. Mercuri, F. Muntoni, B. Sepodes, E. Vroom, P. Balabanov, Development of Exon Skipping Therapies for Duchenne Muscular Dystrophy: A Critical Review and a Perspective on the Outstanding Issues. Nucleic. Acid. Ther. 27(5), 251–259 (2017).

12. A. Aartsma-Rus, M. Bremmer-Bout, A. A. Janson, J. T. den Dunnen, G. J. van Ommen, J. C. van Deutekom, Targeted exon skipping as a potential gene correction therapy for Duchenne muscular dystrophy. Neuromuscul. Disord. 12 Suppl 1, S71–S77 (2002).

13. L. Echevarría, P. Aupy, A. Goyenvalle, Exon-skipping advances for Duchenne muscular dystrophy. Hum. Mol. Genet. 27(R2), R163–R172 (2018).

14. G. C. Kendall, E. I. Mokhonova, M. Moran, N. E. Sejbuk, D. W. Wang, O. Silva, R. T. Wang, L. Martinez, Q. L. Lu, R. Damoiseaux, M. J. Spencer, S. F. Nelson, M. C. Miceli, Dantrolene enhances antisense-mediated exon skipping in human and mouse models of Duchenne muscular dystrophy. Sci. Transl. Med. 4(164), 164ra160 (2012).

15. L. L. Qi, A. Rabinowitz, C. C. Yun, T. Yokota, H. Yin, J. Alter, A. Jadoon, G. Bou-Gharios, T. Partridge, Systemic delivery of antisense oligoribonucleotide restores dystrophin expression in body-wide skeletal muscles. Proc. Natl. Acad. Sci. U. S. A. 102(1), 198–203 (2005).

16. J. Alter, F. Lou, A. Rabinowitz, H. Yin, J. Rosenfeld, S. D Wilton, T. A. Partridge, Q. L. Lu, Systemic delivery of morpholino oligonucleotide restores dystrophin expression bodywide and improves dystrophic pathology. Nat. Med. 12(2), 175–177 (2006).

17. T. Yokota, Q. L. Lu, T. Partridge, M. Kobayashi, A. Nakamura, S. Takeda, E. Hoffman, Efficacy of systemic morpholino exon-skipping in Duchenne dystrophy dogs. Ann. Neurol. 65(6), 667–676 (2009).

18. Y. Aoki, A. Nakamura, T. Yokota, T. Saito, H. Okazawa, T. Nagata, S. Takeda, In-frame dystrophin following exon 51-skipping improves muscle pathology and function in the exon 52-deficient mdx mouse. Mol. Ther. 18(11), 1995–2005 (2010).

19. L. Echevarría, P. Aupy, A. Goyenvalle, Exon-skipping advances for Duchenne muscular dystrophy. Hum. Mol. Genet. 27(R2), R163–R172 (2018).

20. A. Nakamura. Moving towards successful exon-skipping therapy for Duchenne muscular dystrophy. J. Hum. Genet. 62(10), 871–876 (2017).

21. C. L. Bladen, D. Salgado, S. Monges, M. E. Foncuberta, K. Kekou, K. Kosma, H. Dawkins, L. Lamont, A. J. Roy, T. Chamova, V. Guergueltcheva, S. Chan, L. Korngut, C. Campbell, Y. Dai, J. Wang, N. Barišić, P. Brabec, J. Lahdetie, M. C. Walter, O. Schreiber-Katz, V. Karcagi, M. Garami, V. Viswanathan, F. Bayat, F. Buccella, E. Kimura, Z. Koeks, J. C. van den Bergen, M. Rodrigues, R. Roxburgh, A. Lusakowska, A. Kostera-Pruszczyk, J. Zimowski, R. Santos, E. Neagu, S. Artemieva, V. M. Rasic, D. Vojinovic, M. Posada, C. Bloetzer, PY. Jeannet, F. Joncourt, J. Díaz-Manera, E. Gallardo, A. A. Karaduman, H. Topaloğlu, R. El. Sherif, A. Stringer, A. V. Shatillo, A. S. Martin, H. L. Peay, M. I. Bellgard, J. Kirschner, K. M. Flanigan, V. Straub, K. Bushby, J. Verschuuren, A. Aartsma-Rus, C. Béroud, H. Lochmüller, The TREAT-NMD DMD Global Database: analysis of more than 7,000 Duchenne muscular dystrophy mutations. Hum. Mutat. 36(4), 395–402 (2015).

22. Y. Echigoya, Y. Aoki, B. Miskew, D. Panesar, A. Touznik, T. Nagata, J. Tanihata, A. Nakamura, K. Nagaraju, T. Yokota, Long-term efficacy of systemic multiexon skipping targeting dystrophin exons 45-55 with a cocktail of vivo-morpholinos in mdx52 mice. Mol. Ther. Nucleic. Acids. 4(2), e225 (2015).

23. Y. Aoki, M. J. A. Wood. Emerging Oligonucleotide Therapeutics for Rare Neuromuscular Diseases. J. Neuromuscul. Dis. 8(6), 869–884 (2021).

24. S. Cirak, V. Arechavala-Gomeza, M. Guglieri, L. Feng, S. Torelli, K. Anthony, S. Abbs, M. E. Garralda, J. Bourke, D. J. Wells, G. Dickson, M. J. A. Wood, S. D. Wilton, V. Straub, R. Kole, S. B. Shrewsbury, C. Sewry, J. E. Morgan, K. Bushby, F. Muntoni, Exon skipping and dystrophin restoration in patients with Duchenne muscular dystrophy after systemic phosphorodiamidate morpholino oligomer treatment: An open-label, phase 2, dose-escalation study. Lancet. 378(9791), 595–605 (2011).

25. J. R. Mendell, L. R. Rodino-Klapac, Z. Sahenk, K. Roush, L. Bird, L. P. Lowes, L. Alfano, A. M. Gomez, S. Lewis, J. Kota, V. Malik, K. Shontz, C. M. Walker, K. M. Flanigan, M. Corridore, J. R. Kean, H. D. Allen, C. Shilling, K. R. Melia, P. Sazani, J. B. Saoud, E. M. Kaye; Eteplirsen Study Group, Eteplirsen for the treatment of Duchenne muscular dystrophy. Ann. Neurol. 74(5), 637–647 (2013).

26. C. M. McDonald, P. B. Shieh, H. Z. Abdel-Hamid, A. M. Connolly, E. Ciafaloni, K. R. Wagner, N. Goemans, E. Mercuri, N. Khan, E. Koenig, J. Malhotra, W. Zhang, B. Han, J. R. Mendell; the Italian DMD Telethon Registry Study Group, Leuven NMRC Registry Investigators, CINRG Duchenne Natural History Investigators, and PROMOVI Trial Clinical Investigators, Open-Label Evaluation of Eteplirsen in Patients with Duchenne Muscular Dystrophy Amenable to Exon 51 Skipping: PROMOVI Trial. J. Neuromuscul. Dis. 8(6), 989–1001 (2021).

27. D. E. Frank, F. J. Schnell, C. Akana, S. H. El-Husayni, C. A. Desjardins, J. Morgan, J. S. Charleston, V. Sardone, J. Domingos, G. Dickson, V. Straub, M. Guglieri, E. Mercuri, L. Servais, F. Muntoni; SKIP-NMD Study Group. Increased dystrophin production with golodirsen in patients with Duchenne muscular dystrophy. Neurology. 94(21), e2270–e2282 (2020).

28. L. Servais, E. Mercuri, V. Straub, M. Guglieri, A. M. Seferian, M. Scoto, D. Leone, E. Koenig, N. Khan, A. Dugar, X. Wang, B. Han, D. Wang, F. Muntoni; SKIP-NMD Study Group, Long-Term Safety and Efficacy Data of Golodirsen in Ambulatory Patients with Duchenne Muscular Dystrophy Amenable to Exon 53 Skipping: A First-in-human, Multicenter, Two-Part, Open-Label, Phase 1/2 Trial. Nucleic. Acid. Ther. 32(1), 29–39 (2022).

29. A. Aartsma-Rus, D. R. Corey, The 10th Oligonucleotide Therapy Approved: Golodirsen for Duchenne Muscular Dystrophy. Nucleic. Acid. Ther. 30(2), 67–70 (2020).

30. H. Komaki, Y. Takeshima, T. Matsumura, S. Ozasa, M. Funato, E. Takeshita, Y. Iwata, H. Yajima, Y. Egawa, T. Toramoto, M. Tajima, S. Takeda, Viltolarsen in Japanese Duchenne muscular dystrophy patients: A phase 1/2 study. Ann. Clin. Transl. Neurol. 7(12), 2393–2408 (2020).

31. P. R. Clemens, V. K. Rao, A. M. Connolly, A. D. Harper, J. K. Mah, C. M. McDonald, E. C. Smith, C. M. Zaidman, T. Nakagawa; CINRG DNHS Investigators; E. P. Hoffman, Long-Term Functional Efficacy and Safety of Viltolarsen in Patients with Duchenne Muscular Dystrophy. J. Neuromuscul. Dis. 9(4), 493–501 (2022).

32. R. R. Roshmi, T. Yokota. Pharmacological Profile of Viltolarsen for the Treatment of Duchenne Muscular Dystrophy: A Japanese Experience. Clin. Pharmacol. 13, 235–242 (2021).

33. Y. Aoki, T. Nagata, T. Yokota, A. Nakamura, M. J. A. Wood, T. Partridge, S. Takeda, Highly efficient in vivo delivery of PMO into regenerating myotubes and rescue in laminin-α2 chain-null congenital muscular dystrophy mice. Hum. Mol. Genet. 22(24), 4914–4928 (2013).

34. A. R. Findlay, N. Wein, Y. Kaminoh, L. E. Taylor, D. M. Dunn, J. R. Mendell, W. M. King, A. Pestronk, J. M. Florence, K. D. Mathews, R. S. Finkel, K. J. Swoboda, M. T. Howard, J. W. Day, C. McDonald, A. Nicolas, E. L. Rumeur, R. B. Weiss, K. M. Flanigan; United Dystrophinopathy Project, Clinical phenotypes as predictors of the outcome of skipping around DMD exon 45. Ann. Neurol. 77(4), 668–674 (2015).

35. S. D. Wilton, A. M. Fall, P. L. Harding, G. McClorey, C. Coleman, S. Fletcher, Antisense oligonucleotide-induced exon skipping across the human dystrophin gene transcript. Mol. Ther. 15(7), 1288–1296 (2007).

36. N. Watanabe, Y. Tone, T. Nagata, S. Masuda, T. Saito, N. Motohashi, K. Takagaki, Y. Aoki, S. Takeda, Exon 44 skipping in Duchenne muscular dystrophy: NS-089/NCNP-02, a dual-targeting antisense oligonucleotide. Mol. Ther. Nucleic. Acids. 34, 102034 (2023).

37. H. Takizawa, Y. Hara, Y. Mizobe, T. Ohno, S. Suzuki, K. Inoue, E. Takeshita, Y. Shimizu-Motohashi, A. Ishiyama, M. Hoshino, H. Komaki, S. Takeda, Y. Aoki, Modelling Duchenne muscular dystrophy in MYOD1-converted urine-derived cells treated with 3-deazaneplanocin A hydrochloride. Sci. Rep. 9(1), 3807 (2019).

38. H. Takizawa, E. Takeshita, M. Sato, Y. Shimizu-Motohashi, A. Ishiyama, M. Mori-Yoshimura, Y. Takahashi, H. Komaki, Y. Aoki, Highly sensitive screening of antisense sequences for different types of DMD mutations in patients’ urine-derived cells. J. Neurol. Sci. 423, 117337 (2021).

39. Y. Hathout, R. L. Marathi, S. Rayavarapu, A. Zhang, K. J. Brown, H. Seol, H. Gordish-Dressman, S. Cirak, L. Bello, K. Nagaraju, T. Partridge, E. P. Hoffman, S. Takeda, J. K. Mah, E. Henricson, C. McDonald, Discovery of serum protein biomarkers in the mdx mouse model and crossspecies comparison to Duchenne muscular dystrophy patients. Hum. Mol. Genet. 23, 6458–6469 (2014).

40. T. Misaka, A. Yoshihisa, Y. Takeishi, Titin in muscular dystrophy and cardiomyopathy: urinary titin as a novel marker. Clin. Chim. Acta. 495, 123–128 (2019).

41. H. Komaki, T. Nagata, T. Saito, S. Masuda, E. Takeshita, M. Sasaki, H. Tachimori, H. Nakamura, Y. Aoki, S. Takeda, Systemic administration of the antisense oligonucleotide NS-065/NCNP-01 for skipping of exon 53 in patients with Duchenne muscular dystrophy. Sci. Transl. Med. 10(437), eaan0713 (2018).

42. P. R. Clemens, V. K. Rao, A. M. Connolly, A. D. Harper, J. K. Mah, E. C. Smith, C. M. McDonald, C. M. Zaidman, L. P. Morgenroth, H. Osaki, Y. Satou, T. Yamashita, E. P. Hoffman; CINRG DNHS Investigators, Safety, Tolerability, and Efficacy of Viltolarsen in Boys With Duchenne Muscular Dystrophy Amenable to Exon 53 Skipping: A Phase 2 Randomised Clinical Trial. JAMA. Neurol. 77(8), 982–991 (2020).

43. E. P. Hoffman, L. M. Kunkel, C. Angelini, A. Clarke, M. Johnson, J. B. Harris, Improved diagnosis of Becker muscular dystrophy by dystrophin testing. Neurology. 39(8), 1011–1017 (1989).

44. M. Neri, S. Torelli, S. Brown, I. Ugo, P. Sabatelli, L. Merlini, P. Spitali, P. Rimessi, F. Gualandi, C. Sewry, A. Ferlini, F. Muntoni, Dystrophin levels as low as 30% are sufficient to avoid muscular dystrophy in the human. Neuromuscul. Disord. 17(11-12), 913–918 (2007).

45. L. Servais, E. Mercuri, V. Straub, M. Guglieri, A. M. Seferian, M. Scoto, D. Leone, E. Koenig, N. Khan, A. Dugar, X. Wang, B. Han, D. Wang, F. Muntoni; SKIP-NMD Study Group, Long-Term Safety and Efficacy Data of Golodirsen in Ambulatory Patients with Duchenne Muscular Dystrophy Amenable to Exon 53 Skipping: A First-in-human, Multicenter, Two-Part, Open-Label, Phase 1/2 Trial. Nucleic. Acid. Ther. 32(1), 29–39 (2022).

46. C. Brogna, G. Coratti, M. Pane, V. Ricotti, S. Messina, A. D’Amico, C. Bruno, G. Vita, A. Berardinelli, E. Mazzone, F. Magri, F. Ricci, T. Mongini, R. Battini, L. Bello, E. Pegoraro, G. Baranello, S. C. Previtali, L. Politano, G. P. Comi, V. A. Sansone, A. Donati, E. Bertini, F. Muntoni, N. Goemans, E. Mercuri; on behalf on the International DMD group, Long-term natural history data in Duchenne muscular dystrophy ambulant patients with mutations amenable to skip exons 44, 45, 51 and 53. PLoS. One. 14(6), e0218683 (2019).

47. K. Anthony, V. Arechavala-Gomeza, V. Ricotti, S. Torelli, L. Feng, N. Janghra, G. Tasca, M. Guglieri, R. Barresi, A. Armaroli, A. Ferlini, K. Bushby, V. Straub, E. Ricci, C. Sewry, J. Morgan, F. Muntoni, Biochemical characterisation of patients with in-frame or out-of-frame DMD deletions pertinent to exon 44 or 45 skipping. JAMA. Neurol. 71(1), 32–40 (2014).

48. R. C-G. Odel, M. Rodriguez-Cruz, R. E. E. Cedillo, Systemic inflammation in Duchenne muscular dystrophy: association with muscle function and nutritional status. Biomed. Res. Int. 2015, 891972 (2015).

49. H. A. John, I. F. Purdom, Elevated plasma levels of haptoglobin in Duchenne muscular dystrophy:electrophoretic variants in patients with a severe form of the disease. Electrophoresis. 10, 489–493 (1989).

50. V. D. Nadarajah, M. van Putten, A. Chaouch, P. Garrood, V. Straub, H. Lochmu ller, H. B. Ginjaar, A. M. Aartsma-Rus, G. J. B. van Ommen, J. T. den Dunnen, P. A. ’t Hoen, Serum matrix metalloproteinase-9 (MMP-9) as a biomarker for monitoring disease progression in Duchenne muscular dystrophy (DMD). Neuromuscul. Disord. 21, 569–578 (2011).

51. U. J. Dang, M. Ziemba, P. R. Clemens, Y. Hathout, L. S. Conklin, CINRG Vamorolone 002/003 Investigators, E. P. Hoffman, Serum biomarkers associated with baseline clinical severity in young steroid-naïve Duchenne muscular dystrophy boys. Hum. Mol. Genet. 29(15), 2481–2495 (2020).

52. J. Lange, O. Gillham, R. Alkharji, S. Eaton, G. Ferrari, M. Madej, M. Flower, F. S. Tedesco, F. Muntoni, P. Ferretti, Dystrophin deficiency affects human astrocyte properties and response to damage. Glia. 70(3), 466–490 (2022).

53. M. E. Benny Klimek, M. C. Vila, K. Edwards, J. Boehler, J. Novak, A. Zhang, J. V. der Meulen, K. Tatum, J. Quinn, A. Fiorillo, U. Burki, V. Straub, Q. L. Lu, Y. Hathout, J. van D. Anker, T. A. Partridge, M. Morales, E. Hoffman, K. Nagaraju, Effects of Chronic, Maximal Phosphorodiamidate Morpholino Oligomer (PMO) Dosing on Muscle Function and Dystrophin Restoration in a Mouse Model of Duchenne Muscular Dystrophy. J. Neuromuscul. Dis. 8(s2), S369–S381 (2021).

54. T. Ishizuka, H. Komaki, Y. Asahina, H. Nakamura, N. Motohashi, E. Takeshita, Y. Shimizu-Motohashi, A. Ishiyama, C. Yonee, S. Maruyama, E. Hida, Y. Aoki, Systemic administration of the antisense oligonucleotide NS-089/NCNP-02 for skipping of exon 44 in patients with Duchenne muscular dystrophy: study protocol for a phase I/II clinical trial. Neuropsychopharmacol. Rep. 43(2), 277–286 (2023).

55. M. Takase, H. Sakata, M. Shikada, K. Tatara, T. Fukushima, T. Miyakawa, Development of reference equations for spirometry in japanese children aged 6–18 years. Pediatr. Pulmonol. 48(1), 35–44 (2013).

56. F. Muntoni, J. Signorovitch, G. Sajeev, H. Lane, M. Jenkins, I. Dieye, S. J. Ward, C. McDonald, N. Goemans, E. H. Niks, B. Wong, L. Servais, V. Straub, M. Guglieri, I. J. M. de Groot, M. Chesshyre, C. Tian, A. Y. Manzur, E. Mercuri, A. Aartsma-Rus; Association Française Contre Les Myopathies; on behalf of Universitaire Ziekenhuizen Leuven Group, PRO-DMD-01, The UK NorthStar Clinical Network, CCHMC, and The DMD Italian Group, DMD Genotypes and Motor Function in Duchenne Muscular Dystrophy: A Multi-institution Meta-analysis With Implications for Clinical Trials. Neurology. 100(15), e1540–e1554 (2023).

