## Supplemental Information for "Phase I/II Trial of Brogidirsen: Dual-Targeting Antisense Oligonucleotides for Exon 44 Skipping in Duchenne Muscular Dystrophy"

### 1 Department of Child Neurology, National Center Hospital, National Center of Neurology and Psychiatry (NCNP); Kodaira, Tokyo 187-8502, Japan.

### 2 Translational Medical Center, National Center of Neurology and Psychiatry (NCNP); Kodaira, Tokyo 187-8502, Japan.

### 3 Department of Molecular Therapy, National Institute of Neuroscience, National Center of Neurology and Psychiatry (NCNP); Kodaira, Tokyo 187-8502, Japan.

### 4 Department of Pediatrics, Graduate School of Medical and Dental Sciences, Kagoshima University; Kagoshima City, Kagoshima 890-8520, Japan.

### 5 Department of Biostatistics and Data Science, Graduate School of Medicine, Osaka University; Suita-Shi, Osaka 565-0871, Japan.

### **Text S1: Full list of study exclusion criteria**

The full list of exclusion criteria is as follows:

- Participation in another clinical trial designed to restore the expression of dystrophin protein
- Had received another investigational drug in the 3 months prior to initiating NS-089/NCNP-02, brogidirsen
- Impaired respiratory function requiring the continuous use of a ventilator
- Less than 50% of the predicted value (based on the reference values for spirometry in Japanese children) for forced vital capacity
- Less than 40% left ventricular ejection fraction or <25% fractional shortening (by electrocardiogram)
- Immunodeficiency, active uncontrolled infection, autoimmune disease, cardiomyopathy, renal or hepatic disease, were positive for hepatitis B virus surface antigen, hepatitis C virus antibody, or human immunodeficiency virus antibody
- Surgery in the 3 months prior to enrollment or had a planned surgery
- History of serious hypersensitivity reactions to pharmacologic drugs
- Unable to comply with the study procedures because of cognitive challenges

Patient's safety could have been compromised by study participation

**Figure S1:** **Changes in plasma creatinine kinase throughout brogidirsen treatment during**

**Part 2.** Cohort 1 was treated with 40 mg/kg brogidirsen, and Cohort 2 was treated with 80 mg/kg

brogidirsen (administered intravenously once weekly for 24 weeks). CK=creatinine kinase.

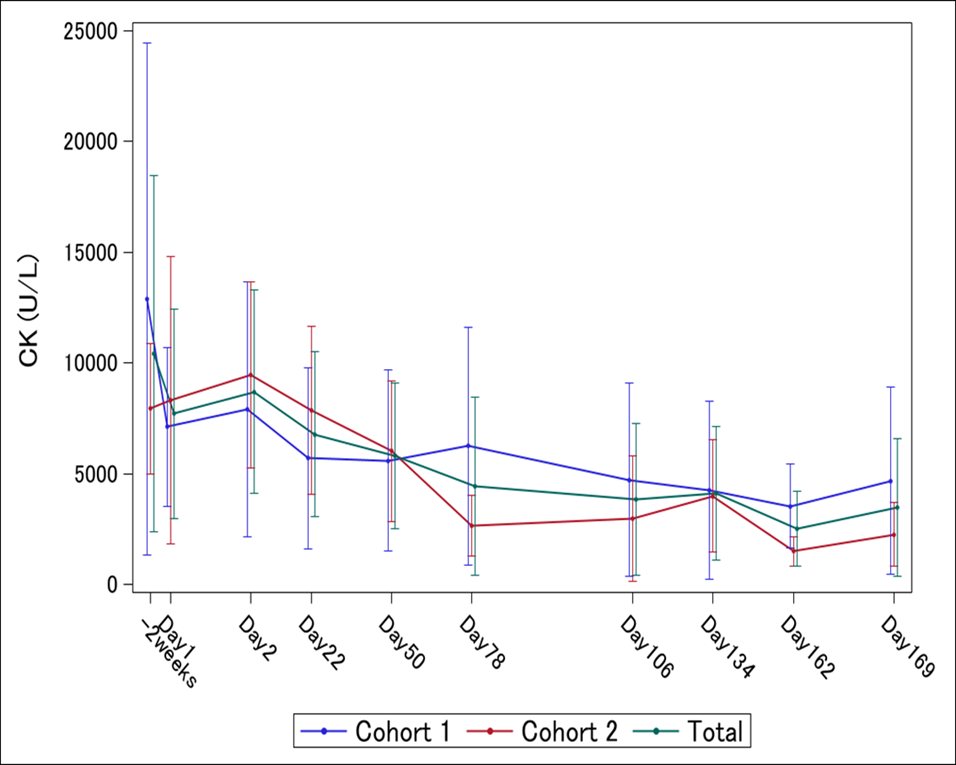

### **Figure S2: Changes of proteomics in plasma during treatment (A) temporal profile of Dystrophin, (B) IL6, (C) TNF, (D) IFNG, (E) SPP1 (osteopontin) and (F) MMP9.** Plasma was collected from each patient at 3 different time points, including 1 week, 12 weeks, and 24 weeks in Part 2. Protein expression was evaluated using proximity extension assays (Olink).

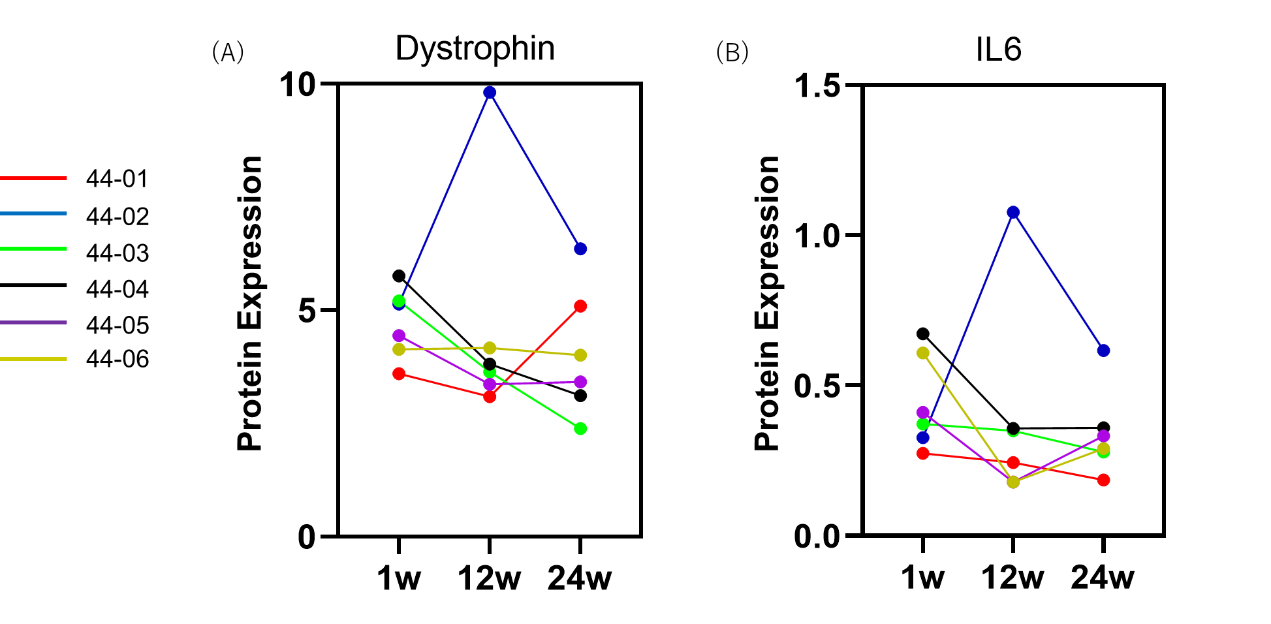

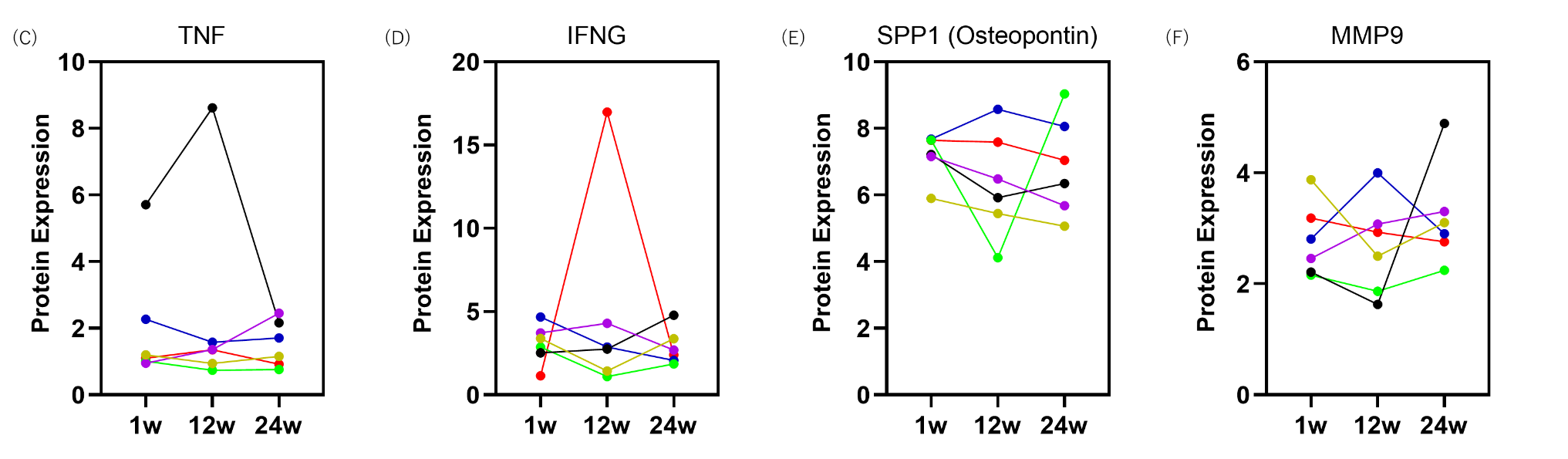

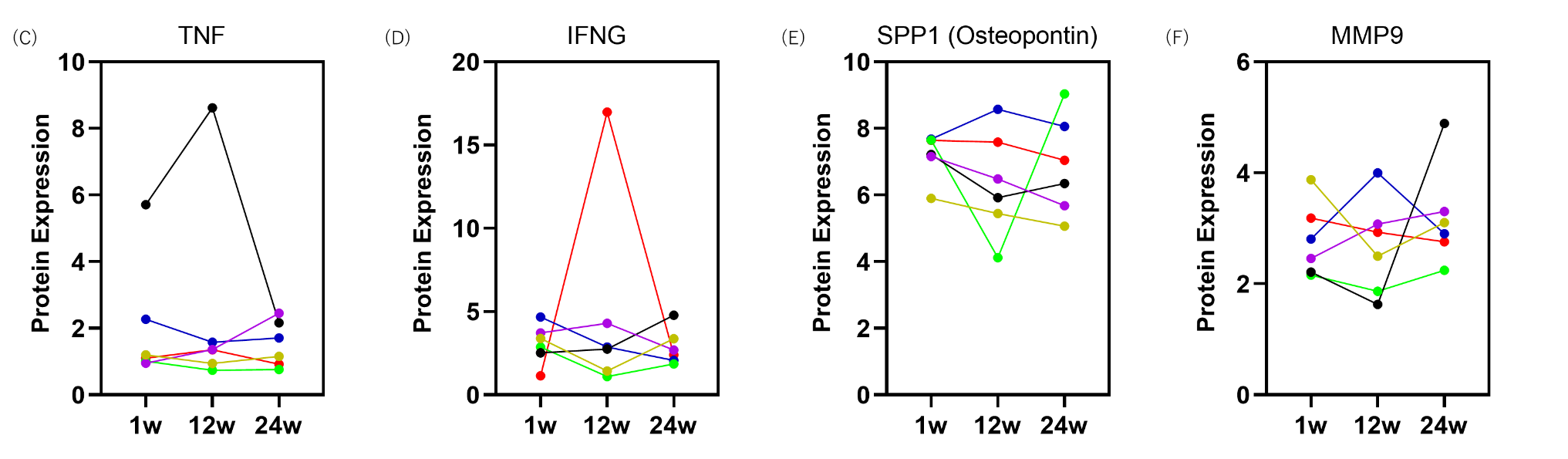

### **Figure S3: Evaluation of (A) exon 44 skipping and dystrophin protein expression in patient-derived myoblast-derived myotubes and (B) patient-derived MYOD1 fibroblasts following *in vitro* treatment with brogidirsen**

(A)

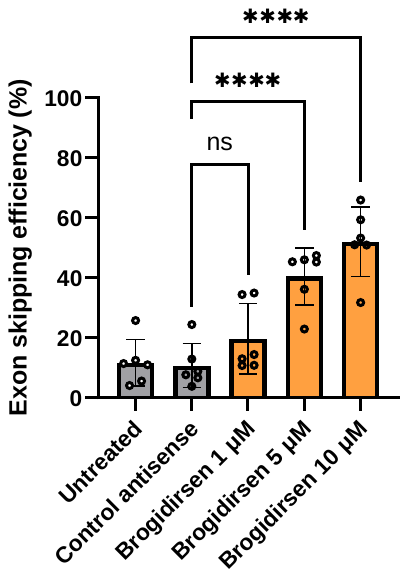

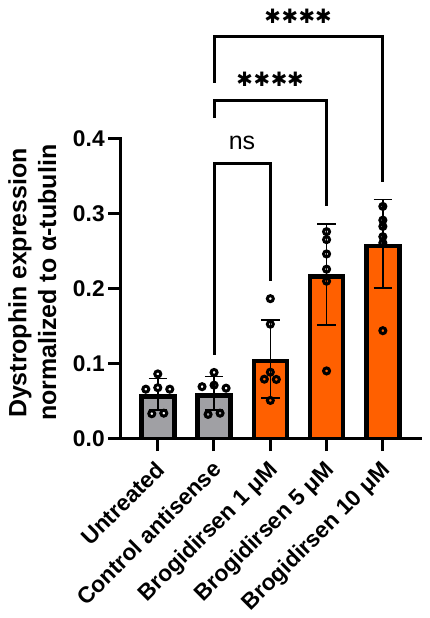

(B)

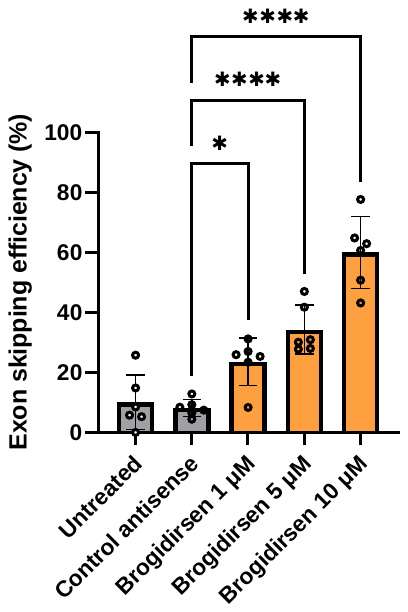

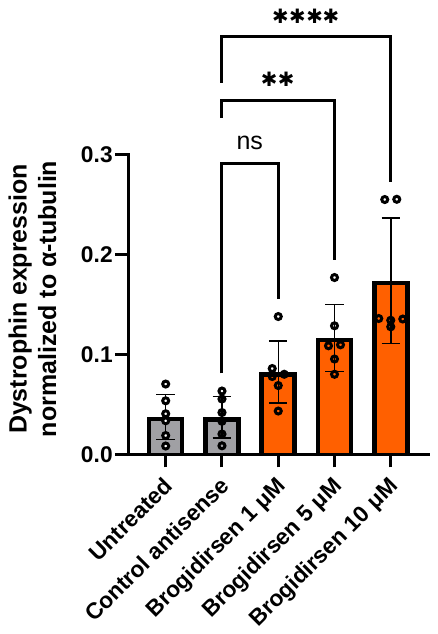

**Figure S4: Study design**

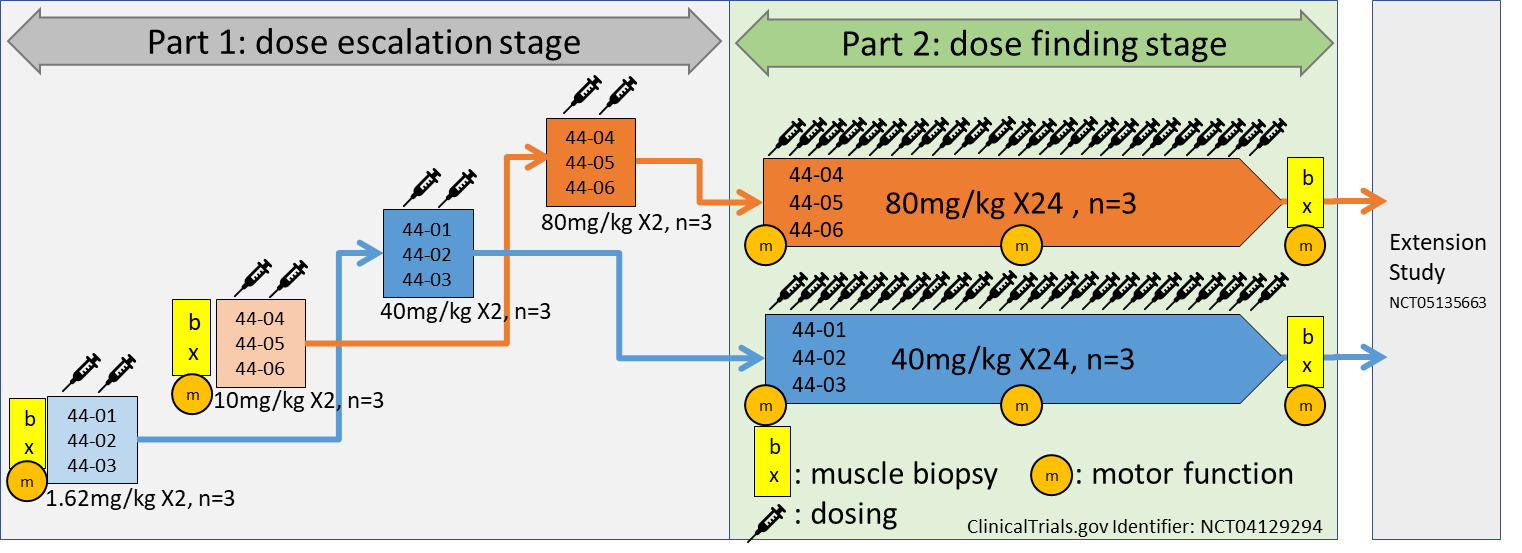

**Table S1: List of primers**

| **Reverse Transcription Polymerase Chain Reaction (RT-PCR) Primers** | | |
| --- | --- | --- |
| **For Tissues or Cells?** | **Primer Name (Mutation Type)** | **Sequence (5'-3')** |
| Tissues | Del. Exon 45_Forward | GCTCAGGTCGGATTGACATT |
|  | Del. Exon 45_Reverse | GGGCAACTCTTCCACCAGTA |
| Tissues | Del. Exon 45-54_Forward | ACAAAGCTCAGGTCGGATTG |
|  | Del. Exon 45-54_Reverse | CCTTGGAGGTCTTGCCATTG |
| Cells | Del. Exon 45_Forward | GCTCAGGTCGGATTGACATT |
|  | Del. Exon 45_Reverse | GGGCAACTCTTCCACCAGTA |
| Cells | Del. Exon 45-54_Forward | ATGCTCCTGACCTCTGTGCT |
|  | Del. Exon 45-54_Reverse | GACTGCATCATCGGAACCTT |
